# Sex and insulin resistance biomarker modelling in a new large-scale Alzheimer’s disease transcriptomic resource

**DOI:** 10.1101/2025.10.02.25337067

**Authors:** Nasim Mohamed Ismail, Maggie A. Miller, Hannah Crossland, Bethan E Phillips, Jalil-Ahmad Sharif, Christina Kosanovic, Tianjie Gu, Robert J Brogan, Claes Wahlestedt, Philip J Atherton, J Paul Chapple, William E Kraus, Kirill Shkura, Anthony J Griswold, Claude-Henry Volmar, Gregory Slabaugh, James A. Timmons

## Abstract

**Background:** The incidence of Alzheimer’s disease (AD) increases with age, is associated with insulin resistance (IR), and has a greater prevalence in women. Genome-wide technologies can yield novel biomarkers and may help identify aspects of AD pathophysiology. Individual AD blood transcriptomic studies are small, preventing the study of sex, while meta-analyses are technically challenging. Relationships between AD biomarkers and IR are also underexplored, largely due to a lack of metabolic phenotyping in AD cohorts.

**Methods:** We generated 1,021 new whole-blood transcriptomic profiles from the AddNeuroMed cohort (including 317 technical replicates) and 410 whole-blood transcript profiles (metabolic cohort). We aligned this data, and our large AD RNA-seq whole-blood transcriptome study, to a common genomic and transcriptomic reference and modelled blood cell composition. Bias was assessed using randomly sampled gene-sets and cross-validated classifiers. Further, a 62-gene IR signature was generated using our metabolic cohort studies, providing a surrogate IR RNA score to retrospectively phenotype the AD cohort. Machine learning was used to develop AD classifiers from the data and evaluate multimodal integration with magnetic resonance imaging. Sex-stratified differential expression and pathway analysis were used to explore sex differences in AD.

**Results:** The new data was more robust than the original AddNeuroMed data, with a lower ‘sampling at random’ score (AUC=0.61 vs 0.73-0.79). A novel AD classification signature was cross- and externally evaluated, achieving higher performance in women. Identification of AD-associated disease pathways, in whole-blood, was influenced by variation in blood cell composition. Notably, B-cell pathways were modified in AD (including genes *BLNK* and *MS4A1*), and this was relatively consistent across sexes and ethnicity. Previous reports that mitochondrial-DNA-encoded RNAs were upregulated, and nuclear-encoded mitochondrial transcripts were consistently downregulated, were not substantiated, with only women showing modest evidence for loss of nuclear-encoded mitochondrial transcripts.

**Conclusions:** We provide a large-scale, technically robust blood AD transcriptomic dataset that enhances legacy AD resources. Analysis revealed robust immune signatures and the statistical transfer of a classification signature across technologies and ethnicities. We add to the evidence for a role of altered B-cell biology in AD, while delivering an updateable transcriptomic resource for future machine learning and genomic studies.

## Background

Alzheimer’s disease (AD) is the most prevalent form of dementia, constituting ∼60% of the estimated 55 million dementia cases worldwide [1]. AD is characterised by progressive cognitive impairment, with definitive diagnosis confirmed postmortem by histopathological identification of Amyloid β (Aβ) plaques and hyperphosphorylated tau neurofibrillary tangles [2]. Importantly, AD begins many years prior to clinical diagnosis [3], while established molecular hallmarks of AD provide only a narrow selection of potential biomarkers or drug targets [4]. Molecular profiling of blood and cerebrospinal fluid (CSF) [5,6] has yielded several robust biomarkers for AD status [7,8]. For example, plasma phosphorylated tau^217^ (p-tau^217^) appears comparable with CSF p-tau^217^ for estimating brain Aβ and tau deposition determined by positron emission tomography (PET), providing a minimally invasive, cost-effective option [7] to capture some aspects of AD pathophysiology. AD disease progression is heterogeneous, with molecular features of resilience emerging [9], and there is a greater prevalence and potentially distinct disease pathology of AD in women [10]. These considerations motivate the need for robust genome-wide molecular studies to identify the breadth of underlying molecular events and to better support drug-target discovery and development.

Whole-blood transcriptomics is a minimally invasive genome-wide molecular analysis, with early studies identifying potential pathological events occurring in the initial stages of AD [11]. There is also evidence that blood transcriptomic profiling can track longitudinal changes in cognitive status [12]. Early work studying differentially expressed genes in whole blood from AD patients reported a reduction in mitochondrial nuclear-encoded oxidative phosphorylation (OXPHOS) and induction of mitochondrial-DNA encoded gene expression [11,13]. Preclinical and clinical studies have also linked altered immune pathways to AD pathogenesis [14], motivating investigation of blood-based molecular changes in AD. Nevertheless, shifts in whole blood cell composition may complicate the interpretability of whole-blood transcriptome modelling. For example, in observational studies, low monocyte and eosinophil counts and higher leukocyte and neutrophil counts are associated with increased risk of AD [15]. Associations between neutrophil count and AD may reflect confounding age-related variables [16,17]. Likewise, insulin resistance (IR) [18] and type 2 diabetes are linked to increased risk of AD and dementia [19,20]. Variation in metabolic status between AD cases and controls would influence the biological interpretation of whole-blood transcriptomic findings. Indeed, even tau phosphorylation AD biomarkers are influenced by cardiometabolic health independent of AD status in real-world data [21]. Thus, differences in gene expression between AD and control subjects can be driven by incidental differences in cell populations or comorbidities, and it remains unclear whether reported molecular differences [11,13] are reliable features of AD.

Systematic non-AD-specific differences could also inflate the performance of machine learning (ML) classifiers trained on AD blood molecular data, impacting their relevance in a real-world setting. Research consortia such as the Alzheimer’s Disease Neuroimaging Initiative (ADNI) and AddNeuroMed have generated valuable biological and neuroimaging resources to support the development of AD prognostics and diagnostics. The largest transcriptomic datasets are limited by technical complications [22–24], including >30% of the gene-labels being inaccurate due to an inability to realign probes against the current genome [25]. For AddNeuroMed (also known as ANMerge) [5], the data has unresolvable batch issues that make it unsuitable for *training* an ML diagnostic classifier. Existing individual AD blood transcriptomics resources are limited in size, making the study of sexual dimorphism challenging [10]. AD blood transcriptomics projects [26] have also utilised RNA sequencing [25], and a recent meta-analysis attempted to model the blood transcriptomic relationship with AD status [27]. This was unsuccessful, perhaps in part due to the reliance on distinct genome references for each data source.

To address these various topics, we generate a new and large-scale blood transcriptomic resource from an extended version of the AddNeuroMed project [24] (Figure 1). The new data utilises a high-throughput array technology, where transcript signals are built from multiple probes which can be realigned to the genome, to ensure that the transcript signal remains accurate over time, just like reprocessing raw sequencing data. We illustrate how this new resource can be used to integrate transcriptomics and magnetic resonance imaging (MRI) features to develop prototype multimodal diagnostic models of AD status and evaluate our transcriptomic signatures across technical platforms and distinct ethnicities. We then illustrate how a novel transcriptomic signature for insulin-resistance (IR), can be used to retrospectively phenotype AD cases and cohort-defined control subjects. We reveal novel and consistent whole-blood molecular features of AD, and report the first blood transcriptomic comparison between men and women with AD.

**Figure 1.**
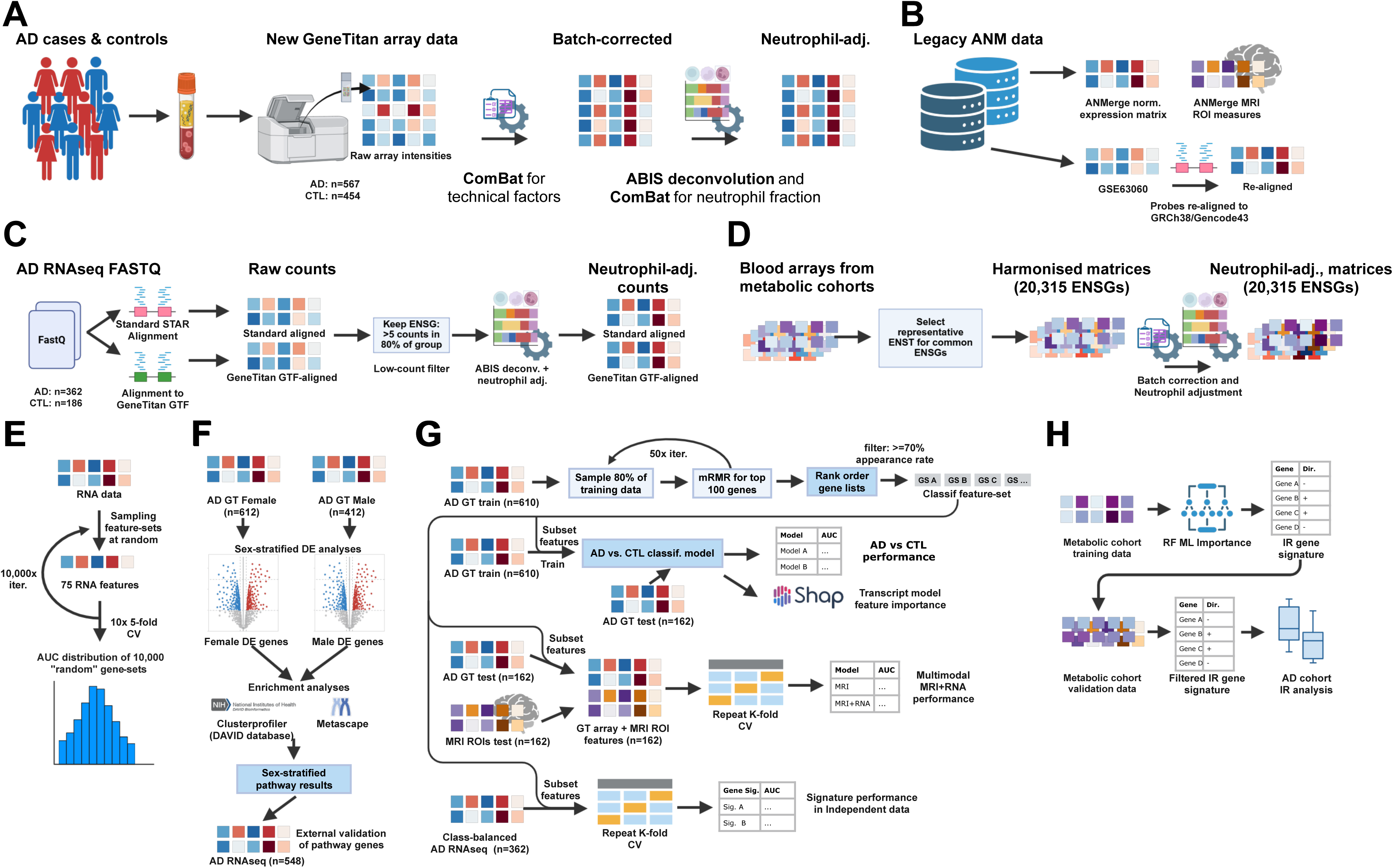
Overview of study design, datasets and modelling strategy. **A:** Generation and preprocessing of the present new AddNeuroMed (ANM) whole-blood Affymetrix GeneTitan transcriptomic dataset, including probe realignment to GRCh38/Gencode43, IRON normalisation, ComBat adjustment for plate and site, and additional correction for neutrophil fraction to yield whole-blood and neutrophil-adjusted matrices. **B:** Processing of legacy Illumina ANM microarray data, including probe realignment, comparison of alternative normalisations (Illumina ANM vs Illumina GSE63060) and integration with MRI ROI measures from the ANMerge resource. **C:** RNA-seq processing pipeline for the independent Miami AD cohort, showing standard STAR alignment versus GeneTitan-GTF-based alignment, count filtering, DESeq2 normalisation, ComBat-SEQ correction for library size and sequential neutrophil adjustment, harmonised to genes detected on the GeneTitan array. **D:** Preprocessing and harmonisation of whole-blood microarray data from three metabolic cohorts (MP, SPD, S2), including batch and sex correction, immune deconvolution and neutrophil adjustment to generate common 20,315-gene matrices. E: Gene set sampling framework used to quantify dataset-specific bias, in which 75-gene sets are sampled at random and evaluated with logistic regression using 5×5-fold cross-validation over 10,000 iterations. **F:** Sex-stratified differential expression analyses in GeneTitan data, with pathway enrichment performed using DAVID and Metascape and supported by RedRibbon rank-based concordance analyses. **G:** Construction and evaluation of AD blood transcriptomic and multimodal (transcriptomics–MRI) classifiers, including propensity-score matching, repeated mRMR-based feature selection, nested cross-validated model training and held-out testing, and SHAP-based estimation of feature importance. **H:** Derivation of a cross-platform insulin resistance (IR) blood transcriptomic signature from metabolic cohorts using random forest feature-importance, filtering for cross-cohort concordance, validation against log2 HOMA2-IR in multiple datasets, and application of the final 62-gene IR signature to the AD GeneTitan cohort. Abbreviations: ANM, AddNeuroMed; MRI, magnetic resonance imaging; ROI, region of interest; MP, META-PREDICT; SPD, STRRIDE-PD; S2, STRRIDE-AT/RT; IR, insulin resistance.

## Methods

### Whole Blood AD cohort

Blood samples from subjects belonging to the AddNeuroMed consortium, a large cross-European AD biomarker study, and the Dementia Case Register (DCR) cohort, were provided to us by Dr Angela Hodges (King’s College London) and processed in our laboratory. As per the original study protocol, samples from subjects were excluded if they had neurological or psychiatric conditions other than AD, demonstrated a Geriatric Depression Scale score ≥ 4/5 or other unstable systemic illness. AD was diagnosed using the National Institute of Neurological and Communicative Disorders and Stroke and Alzheimer’s disease (NINCDS-ADRDA) and Diagnostic and Statistical Manual of Mental Disorders (DSM-IV) criteria for AD. Each subject underwent an interview and neuropsychological assessments (e.g. Mini-Mental State Examination (MMSE)). Controls were also assessed using the CERAD battery [28]. Venous blood was collected into PAXgene™ tubes (Becton & Dickinson, Qiagen Inc., Valencia, CA), which were frozen at -20^◦^C and then stored at -80^◦^C. RNA was extracted using PAXgene™ Blood RNA Kit (Qiagen) according to the manufacturer’s instructions.

### Transcriptomic dataset production, genomic annotation and quality control

A total of 1,021 AD and control RNA samples were successfully profiled (passing standard quality checks), from 371 individuals with an AD diagnosis and 333 controls. We included 317 technical replicates (from 149 AD and 115 controls). RNA was profiled using the high-throughput Affymetrix GeneTitan platform and the HTHGU133Plus PM array, following the manufacturer’s protocols (Karolinska Institute Core Facility, Huddinge, Sweden). Each transcript signal is built from a combination of 25-mer probes, and each one was realigned to genomic and transcriptomic references [25]. Briefly, a FASTA file representing the original array design was aligned against Grch38 - Gencode 43 [29] using the STAR aligner [30], and probes with unique matches were retained. Probes with both a very low signal and a low coefficient of variation were removed as these were considered background signal. The remaining probes were combined to form “probe-sets”, each with at least three probes, into a custom map file, known as a custom CDF. This process can be repeated in the future, ensuring the probes remain accurately aligned to the latest genome. Data was normalised using iterative rank-order normalization (IRON) [31] in the default mode, and subject to standard quality checks [25,32]. The custom CDFs and all new raw data files are deposited at the Array Express (E-MTAB-15140), along with normalised data. In the present study, for the AD data, 59,493 probe-sets (filtered using absolute standard deviation values) [25,32] were considered expressed in whole blood, representing 28,423 probe-sets with distinct signals per gene, reflecting a final total of 11,596 genes (see Supplementary Data Table 1). It is noteworthy that the ADNI consortium [7] used similar technology to the present work, but the raw data (i.e. the CEL files) are no longer accessible, preventing modernisation of those data for integration into the present analyses, as using annotations from over a decade ago would result in >30% of transcripts not targeting the expected gene.

A subset of the samples used to generate the new GeneTitan data was originally profiled using the Illumina Human HT-12 Expression BeadChip arrays. These data (GSE63060 and GSE63061) suffer from a batch effect of unknown origin, segregating cases and controls. The data are therefore not suited for ML classification modelling, nor ideal for testing hypothesis-based gene-expression classifiers [22–24]. We refer to this data as the “Illumina” data in the present article. We checked the validity of the Illumina HT12 V3 probes (these are single longer probes), by aligning their sequences against GRCh38 - Gencode 43 [29] using STAR aligner [30] and invalid probes were discarded (see supplementary methods). The same Illumina data has recently been reprocessed by the ANMerge team [5] and we also used their data (GSE63060), comparing it with our reprocessing of the same data (Supplementary Figure 1). Our reprocessing of the ANMerge Illumina data yielded a greater number of transcripts than reported by the ANMerge portal (See Table 1 for an overview of data resources). We distinguish between these two distinctly processed data as “Illumina ANM” and “Illumina GSE63060” in this study. The genes detected using the Illumina platforms are listed in Supplementary Data Table 1.

**Table 1.** Summary statistics for samples in the new GeneTitan transcriptomic dataset and ANMerge Illumina HT12 data.

| **Feature** | **GeneTitan** | **Illumina ANM** |
| --- | --- | --- |
| **Samples** | 1021 | 511 |
| **AD** | 567 | 275 |
| **CTL** | 454 | 236 |
| **Female/Male, %** | 59.65% | 62.21% |
| **Age, mean** | 75.77 | 75.14 |
| **Age, range** | [41,95] | [52,88] |
| **Genes** | 21015 | 5212* |

n = 1,021 GeneTitan profiles passed QC and are reported here and deposited in ArrayExpress (E-MTAB-15140). The Illumina ANMerge dataset includes 511 normalised AD and CTL blood transcriptomes profiled on Illumina HT12 V3 and V4 (excluding MCI profiles), summed across both platforms. AD and CTL counts are based on final clinical diagnosis labels. \*: Number of genes available in normalised Illumina ANM data. In our study, we also realigned and annotated GSE63060 (AddNeuroMed batch 1 Illumina HT12 V3, deposited in Gene Expression Omnibus) raw data, which is largely identical as it comes from the same laboratory files used by ANMerge, to the latest genome in 2024 to acquire data representing 12061 genes. Abbreviations: AD, Alzheimer's disease; CTL, control.

### Adjustment for variation in whole blood cell content

Whole blood is a mixture of cell types, and there are potentially coincidental or non-specific (for AD) shifts [15,33,34] in white blood cell populations between controls and AD subjects. Therefore, we applied cellular deconvolution [17,35] to estimate white blood cell subtypes in each sample and modelled how they may contribute to transcript differences between cases and controls. To adjust the data for major technical factors, including the influence of the most abundant white cell types, we used the ComBat package and supervised batch adjustment [36]. In this case, the IRON [31] normalised gene expression data were adjusted by total plate count (18 different 96-well plates contributed to the new GeneTitan dataset) and the clinical site from which the samples were obtained (modest influence). We refer to this adjusted dataset as the “whole blood” transcriptomic dataset. A further correction was applied to the whole blood data, adjusting for the variation in neutrophil count (the largest subcategory of blood cells, see Supplementary Figure 2 and Results). We refer to this dataset as the ‘neutrophil-adjusted’ transcriptomic dataset.

### Sampling ensemble gene sets to evaluate systematic bias

Earlier work using the original AddNeuroMed Illumina data illustrated a batch effect of unknown origin between AD and control blood profiles [22], complicating its use [24]. The data are not suitable for *building* machine learning classification models. The ANMerge consortium recently reported reprocessing the Illumina data to better control for bias [22]. We provide the first evaluation of the bias in these newly processed data, along with the new GeneTitan data obtained using largely the same samples. We used a logistic regression classification method with default hyperparameters. Performance was evaluated using 5 iterations of 5-fold cross-validation (5x5), repeated over 10,000 iterations with gene sets of n=75 features sampled at random, each selected from the full dataset (Figure 1E, Supplementary Figure 3A). A dataset that lacks any difference between cases and controls should return a classification performance closer to the theoretical AUC = 0.5.

### Differential gene expression and enrichment analysis

Differential gene expression (DE) analysis was performed using Significance Analysis of Microarray (SAM) [37] and all the available AD and control samples. The focus was on comparing the influence of sex and white blood cell content on the biological processes attributed to AD (Figure 1F, Input data reported in Supplementary Table 2). Additionally, due to the limitations of using fixed statistical thresholds for studying the level of agreement between two analyses, we used the RedRibbon R (RR) Package [38]. RR is a rank order method relying on modelling the hypergeometric directional distribution of fold-change values between women (control vs. AD) and men (control vs. AD). The “enrichment zones” within the hypergeometric distribution plots produced by RR (See Results) were analysed using pathway analysis. Gene ontology [39] enrichment was computed using the Metascape database [40] and the DAVID Database (Database for Annotation, Visualization, and Integrated Discovery (DAVID), https://david.ncifcrf.gov/) [39], with p-values generated versus the background of genes estimated to be expressed in our blood samples in both cases [41]. DAVID ‘GOTERM_BP_ALL’ results were processed and visualised using the ClusterProfiler R package [42]. This approach enabled us to contrast the biological pathways associated with AD before and after adjustment for neutrophil counts, revealing the extent to which cell types may contribute to observed transcriptomic signatures.

### ANMerge MRI data with matching transcriptomics

ANMerge [5], a rebranded version of the AddNeuroMed project, provides important multimodal data for integrative AD analysis such as structural MRI, clinical data and several omics modalities, including genomics and plasma proteomics. We propose that the new transcriptomic data herein replace the ANMerge transcriptomics resource. Our transcriptomic data use subject identifiers that match back to the internal ANMerge database (Supplementary Table S3). To illustrate the utility of the ANMerge database for multimodal classification, we used processed structural MRI data acquired at the baseline visit for each subject available from the ANMerge portal, along with our new, more reliable transcriptomics data. The MRI data were acquired with 1.5 Tesla T1-weighted MRI protocols and are available as either raw MRI images or processed features. The processed features represent regional brain volumes and cortical thickness measurements for regions of interest (ROIs) computed using FreeSurfer (version 6.0), a software package for analysing the functional, connectional, and structural properties of human brains using neuroimaging data [5,43]. Where multiple entry rows were present for a subject at their baseline visit, each MRI feature was represented by the mean value across all rows. 4 features with zero variance were dropped, yielding 132 features used to train MRI and transcriptomics-MRI models.

### AD cohort data use and model feature selection

New transcriptomics data were available for 1021 AD and control transcriptomics samples, including technical replicates (from a total of 1,666 samples that were run, including individuals with a mild cognitive impairment (MCI) label). For the binary classification analysis, we excluded 57 samples with moderately elevated normalised unscaled standard error, leaving 964 samples. We have not released the MCI samples due to the ambiguity of their true clinical phenotype with respect to future AD status. We will release the MCI data when this ambiguity can be overcome, and the potential for generating misleading analysis can be avoided. Among the 964 AD and control samples, there were 298 technical replicates, providing a further new and unique resource for future work.

For evaluating classification models, a subset of 162 subjects with matched GeneTitan transcriptomic and FreeSurfer MRI data was designated as the held-out test set. The remaining 733 samples (including replicates from subjects not in the held-out group) were used to create a matched training set for transcriptomics ML models. To limit the influence of sex and age on feature selection, propensity score matching was used to create a matched subset of AD and controls for model training. The Python package psmpy was used to calculate propensity logit values, using the clinical variables sex and age as covariates. The samples used for training transcriptomic models (n=733) were sex and age-matched using psmpy to create class-balanced data (n=610). Summary statistics for the matched training set and transcriptomics-only held-out data are shown in Table 2.

**Table 2.**
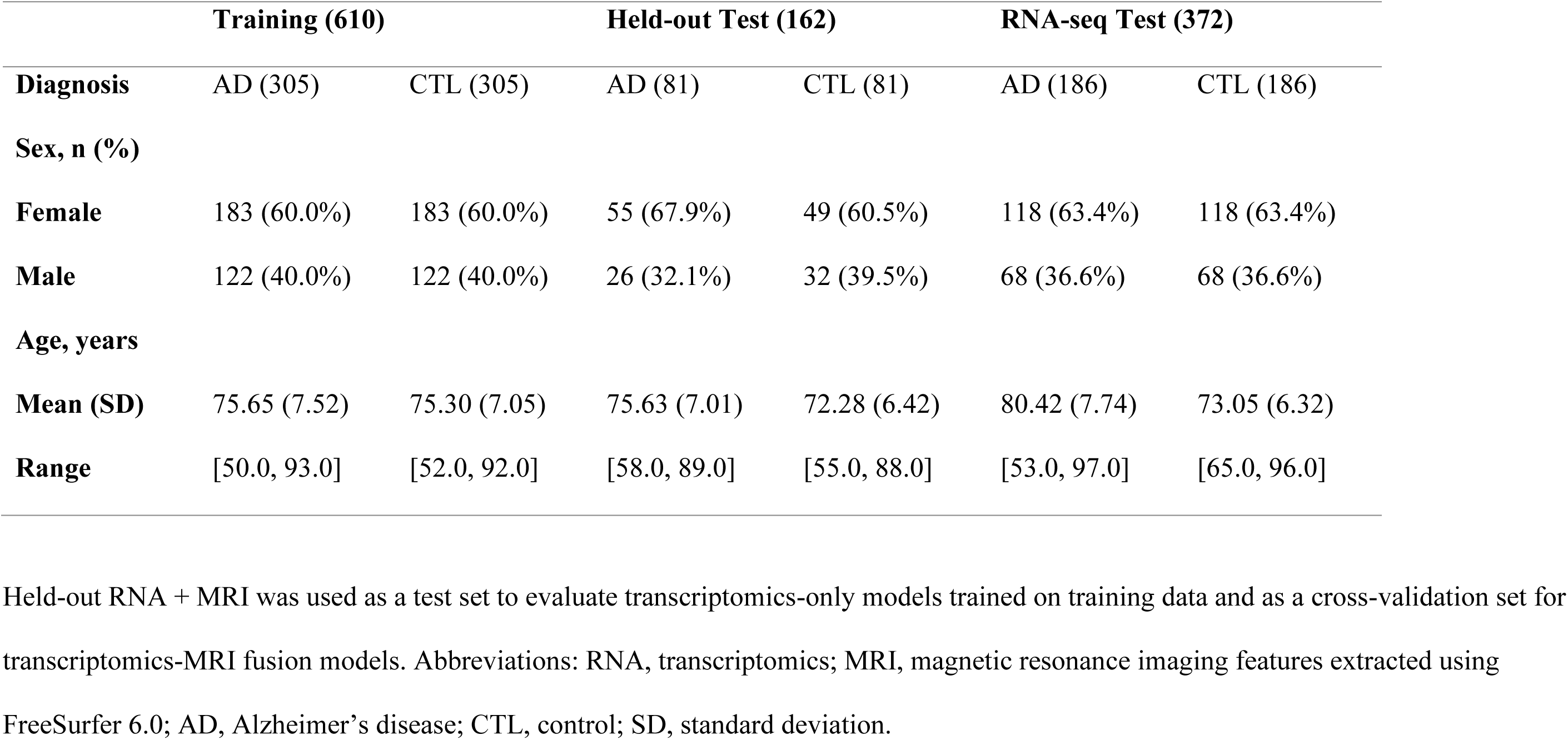
Summary statistics for AD GeneTitan array training and held-out sets, and Grizwold et al. RNA-seq.

|  | **Training (610)** |  | **Held-out Test (162)** |  | **RNA-seq Test (372)** |  |
| --- | --- | --- | --- | --- | --- | --- |
| **Diagnosis** | AD (305) | CTL (305) | AD (81) | CTL (81) | AD (186) | CTL (186) |
| **Sex, n (%)** |  |  |  |  |  |  |
| **Female** | 183 (60.0%) | 183 (60.0%) | 55 (67.9%) | 49 (60.5%) | 118 (63.4%) | 118 (63.4%) |
| **Male** | 122 (40.0%) | 122 (40.0%) | 26 (32.1%) | 32 (39.5%) | 68 (36.6%) | 68 (36.6%) |
| **Age, years** |  |  |  |  |  |  |
| **Mean (SD)** | 75.65 (7.52) | 75.30 (7.05) | 75.63 (7.01) | 72.28 (6.42) | 80.42 (7.74) | 73.05 (6.32) |
| **Range** | [50.0, 93.0] | [52.0, 92.0] | [58.0, 89.0] | [55.0, 88.0] | [53.0, 97.0] | [65.0, 96.0] |

Held-out RNA + MRI was used as a test set to evaluate transcriptomics-only models trained on training data and as a cross-validation set for transcriptomics-MRI fusion models. Abbreviations: RNA, transcriptomics; MRI, magnetic resonance imaging features extracted using FreeSurfer 6.0; AD, Alzheimer’s disease; CTL, control; SD, standard deviation.

Transcriptomics features for model input were selected via a two-step feature selection process using the minimum Redundancy Maximum Relevance (mRMR) method (Figure 1G, Supplementary Figure 3B) [44]. We first applied mRMR to the matched training set to create a list of the top 100 features, which were both highly related to supervised classification labels and as uncorrelated as possible from each other. To minimise the effect of individual samples on mRMR feature selection, we employed a repeated subsampling approach where an 80% subsample of subjects, stratified for diagnostic class (AD or control) and sex (Female or Male), was drawn from the matched training set as input for mRMR, producing 50 ranked lists of the top 100 features per loop. Each transcriptomic feature was then assigned a rank value based on its placement across all lists. If a feature (gene) did not appear in one of the rank order lists, it was assigned a value of 0 for that list. The average rank value for each feature across all 50 rank order lists was calculated to create a final rank order. This consensus rank order list was then filtered to retain genes with an appearance rate of >= 70% across all iterations of mRMR. A strict threshold was chosen to reduce the dimensionality of the AD gene signatures and reduce overfitting, while prioritising the selection of the most discriminant features. This two-stage selection strategy allowed us to balance relevance, non-redundancy, robustness to sample variability, and classifier performance, providing a well-grounded transcriptomic feature set for use in downstream modelling. This is not an exhaustive strategy for deriving an optimal transcriptomic signature, just one that illustrates the potential use of the new transcriptomic resource.

### AD Model training and evaluation

Transcriptomics-only models were trained on the matched training set (n=610) and evaluated on the held-out test set (n=162), with hyperparameter tuning being performed on the training set. For evaluating the performance of MRI and transcriptomics-MRI models, we used 50 repeats of 5-fold cross-validation, stratified for diagnostic class (AD or control) and sex due to a lack of independent MRI data. For each training fold, hyperparameters were optimised exclusively on the training data, using an exhaustive grid search with nested stratified cross-validation. Performance metrics are reported as AUC, sensitivity, specificity, accuracy in females, and accuracy in males. For cross-validated models, metrics were averaged across all test folds. SHAP (SHapley Additive exPlanations) [45] was used to illustrate how each feature influences predictions, capturing both the size and direction of each feature’s contribution to the model’s predictions, based on the mean absolute Shapley value.

### Miami AD cohort and RNA-seq transcriptomic dataset

We evaluated the new transcriptomic classifier signatures in our RNA-seq AD cohort, which included a distinct ethnic composition [26]. FASTQ files were aligned using Star Aligner [30]. To assess the impact of better matching across technologies, an alternate array gene transfer format (GTF)-based alignment was also included. Alignment methods yielded 62,708 ENSGs for standard alignment (very many with zero counts), and 24,481 ENSGs for array GTF-based alignment. Data was filtered to remove genes with less than 5 counts in either the AD or control groups, yielding 16,594 genes for standard aligned data and 13,447 genes for array GTF-aligned data (which is a protein-coding biased array). Count-filtered matrices were then size-factor normalised using DESEQ [46] and log2 transformed. QC visualisation revealed that early principal components were modestly correlated with total counts despite DESEQ2 normalisation. To reduce total count-associated variance, we applied Combat-SEQ [47] to the filtered raw count matrices by binning total counts per sample into decile-based pseudo-batches (Supplementary Figure 4A and 4C). These corrected whole-blood data were then used for cellular deconvolution analysis to produce blood immune fraction estimates and then corrected for the estimated neutrophil fraction (Supplementary Figure 4B and 4D), to produce a neutrophil-adjusted counts matrix. To harmonise gene selection with the GeneTitan AD data for downstream replication analyses, normalised matrices were filtered for genes *detected* in the GeneTitan array experiment (11,596 genes), yielding 10,049 genes from RNA-seq, for the standard aligned data and 9,905 genes for the array GTF-aligned data. Although the GeneTitan GTF defines the full array probe universe, only a subset of these genes passed RNA-seq low-count filtering in our samples, which explains why fewer genes were retained for the array GTF-aligned data.

To assess whether a GeneTitan-derived AD gene expression signature transferred to an RNA-seq-based data set, we performed external validation of two of our binary classification signatures (whole blood and neutrophil-adjusted). We assessed performance using 10 iterations of stratified 5-fold cross-validation, training whole-blood and neutrophil-corrected datasets using genes from the neutrophil-corrected signature. We then compared the performance of the two signatures across different classification algorithms, using the Wilcoxon signed-rank test with FDR correction for multiple testing. We also applied ‘sampling at random’ to the RNA-seq data, to establish the level of systematic bias between cases and controls, to clarify if that data was suitable for building a classification model using internal cross-validation.

### Metabolic cohorts and array transcriptomic datasets

Metabolic disease appears to increase the risk of developing AD [19], and IR has been mechanistically linked to promoting neurodegeneration via altered tau hyperphosphorylation [20]. As metabolic disease is highly prevalent in the general population, it’s also a confounding variable when applying an AD biomarker in real life. IR is not typically measured in neuropsychiatry cohorts, including AddNeuroMed. We therefore developed a novel surrogate IR transcriptomic biomarker to retrospectively phenotype ANMerge/AddNeuroMed. Building on our recent work that developed clinical and metabolomic biomarker models of IR [48], we used new whole-blood transcriptomic data to derive a blood transcriptomic signature discriminating between high and low IR status, segregating a clinically meaningful range [48]. The transcriptomic signature was built using a training set of META-PREDICT [49] (MP) and STRRIDE-PD [50] (SPD) blood profiles (combined N=250; Table 3). Fasting samples from sedentary adults with metabolic risk factors were also profiled using the Affymetrix HTA platform. Transcripts were selected to maximise the number of common transcripts-gene assignments across all metabolic cohort datasets, yielding normalised matrices measuring 20,315 ENSGs (Supplementary Table 4). The data were profiled to compute blood cell estimates using deconvolution [35], and ComBat was used to adjust for variance associated with estimated neutrophil fraction. Visualisation revealed that the datasets showed variance associated with batch variables, and so sequential ComBat correction was applied to reduce the influence of these technical variables (see supplementary filenames for adjusted batch variables). A further adjustment for sex was applied to facilitate the detection of a sex-agnostic IR signature. To test whether the resulting signature generalised across technologies, we filtered for cross-platform concordance and evaluated the signature in the STRRIDE-AT/RT (S2) cohort, profiled on the U133 Plus 2.0 array, and in an additional subset of SPD samples profiled with our AD cohort, on the GeneTitan platform (see sample sizes in Table 3).

**Table 3.**
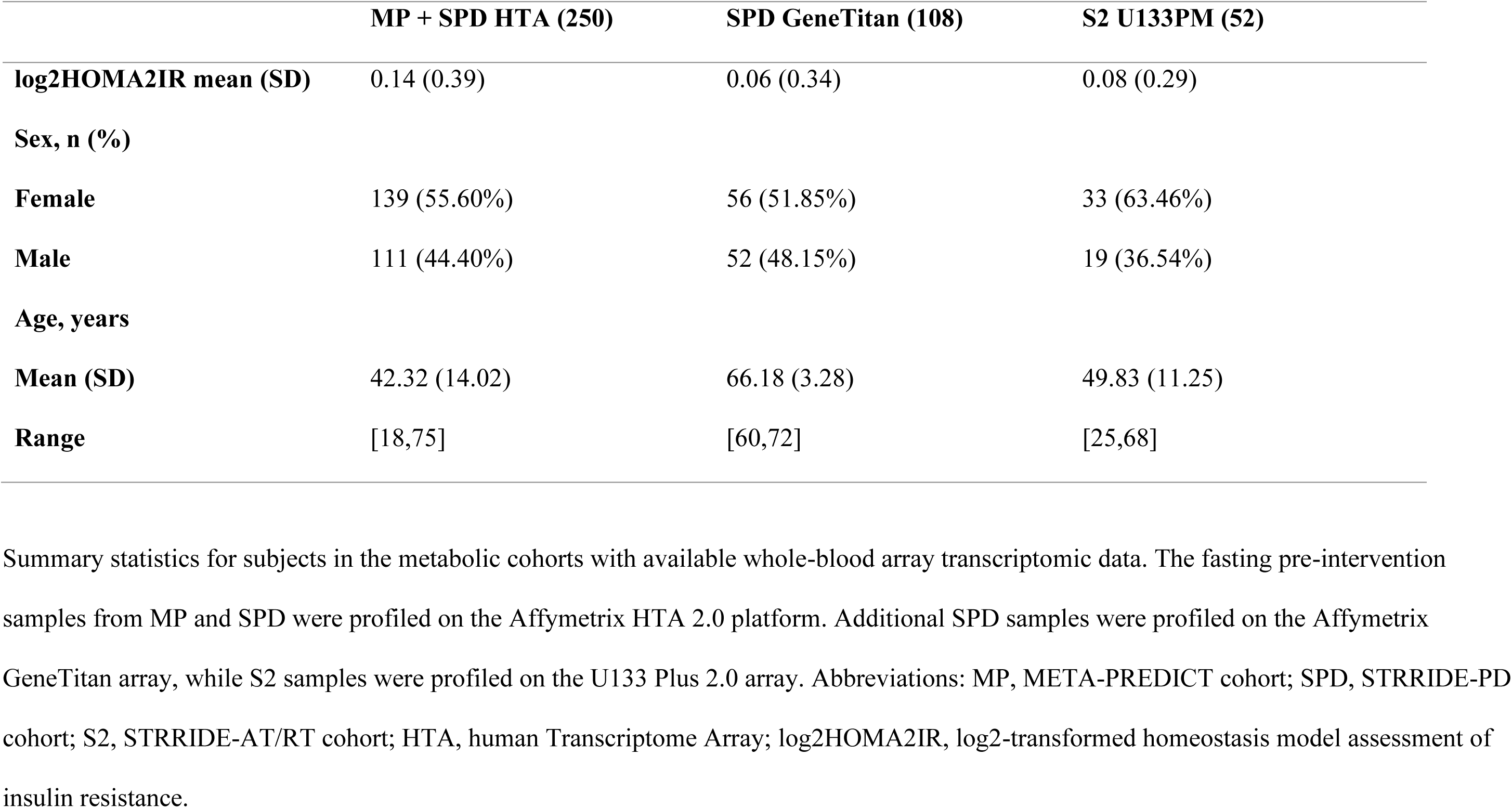
Summary statistics for subjects in blood transcriptomic array datasets in metabolic study cohorts.

|  | **MP + SPD HTA (250)** | **SPD GeneTitan (108)** | **S2 U133PM (52)** |
| --- | --- | --- | --- |
| **log2HOMA2IR mean (SD)** | 0.14 (0.39) | 0.06 (0.34) | 0.08 (0.29) |
| **Sex, n (%)** |  |  |  |
| **Female** | 139 (55.60%) | 56 (51.85%) | 33 (63.46%) |
| **Male** | 111 (44.40%) | 52 (48.15%) | 19 (36.54%) |
| **Age, years** |  |  |  |
| **Mean (SD)** | 42.32 (14.02) | 66.18 (3.28) | 49.83 (11.25) |
| **Range** | [18,75] | [60,72] | [25,68] |

Summary statistics for subjects in the metabolic cohorts with available whole-blood array transcriptomic data. The fasting pre-intervention samples from MP and SPD were profiled on the Affymetrix HTA 2.0 platform. Additional SPD samples were profiled on the Affymetrix GeneTitan array, while S2 samples were profiled on the U133 Plus 2.0 array. Abbreviations: MP, META-PREDICT cohort; SPD, STRRIDE-PD cohort; S2, STRRIDE-AT/RT cohort; HTA, human Transcriptome Array; log2HOMA2IR, log2-transformed homeostasis model assessment of insulin resistance.

### Development of an IR transcriptomic signature and application to the AD cohort

To assess the group-level distribution of IR-associated gene expression in case and controls from the ANMerge/AddNeuroMed GeneTitan cohort, we leveraged model-based feature importance for discriminating between “high” and “low” IR subjects using blood transcriptome profiles (Figure 1H). To build the initial signature, the development set (compromised of profiles from the MP and SPD cohorts profiled on HTA arrays) was subset to the subjects with the top and bottom 25% log2-transformed Homeostatic Model Assessment 2 of Insulin Resistance (HOMA2-IR) values from each cohort[48], resulting in 128 samples matched for high and low IR status. 500 iterations of stratified 10-fold cross-validation splitting was then used, where the training set was used to train a random forest classifier for a binary classification task separating high and low IR profiles. In each split, random forest feature importance values for all 20,315 gene-level features. 500 iterations of 10-fold cross-validation yielded 5000 sets of feature importance values. Gene features with a non-zero importance which appeared in >=20% of all 5000 folds with consistent direction were chosen for the initial signature. These features were used to compute rank gene scores per sample, which were validated by computing the Spearman correlation coefficient between rank gene score and log2 HOMA2IR when the data had a continuous range of values. This evaluation was performed in combined MP+SPD data, the SPD GeneTitan data, and the S2 U133PM data. Rank gene scores generated on all samples from the combined MP and SPD data showed strong correlation with log2 HOMA2IR (Supplementary Figure 5A, Spearman Rho=0.745, *P* < 0.001). This initial signature cleanly separated samples by log2 HOMA2IR status in the SPD GeneTitan data (Supplementary Figure 5B) and in the extremes of the S2 U133PM HOMA2IR values, but not across the entire range (Supplementary Figure 5C, Spearman Rho=0.156, *P*=0.269). Investigating the direction of the univariate correlation between the 147 IR genes revealed that 62/147 (42.2%) genes (Supplementary Table 5) showed concordant direction across all datasets. We therefore relied on only the directionally concordant gene correlations, and 51 genes detected in the AD cohort data were used to compute an IR gene score per sample.

### Statistical analysis

All statistical analyses were performed in R version 4.5.2 with packages samr 3.0.0, DESeq2 1.50.0 and RedRibbon 1.3.0, and in Python 3.11 with scipy 1.17.0 and psmpy 0.3.16, unless otherwise stated. Classification performance was quantified using the area under the receiver operating characteristic curve (AUC). To minimise confounding by age and sex in classifier training, propensity score matching was applied using psmpy to derive a class-balanced subset of AD and control subjects matched on age and sex from the AD GeneTitan data, and transcriptomics-only classifiers were trained on this matched subset. For sampling-at-random experiments, logistic regression classifiers were trained on gene sets sampled at random and evaluated using 10 iterations of 5-fold cross-validation (50 folds total per experiment), repeated over 10,000 iterations; summary AUC values are reported as mean ± standard deviation across iterations. For evaluation of AD blood signatures in the external cohorts, we similarly generated 10,000 random gene sets of equal size to the pre-specified signature, trained the same classification algorithm using fixed default hyperparameter with identical 10×5-fold cross-validation, and used the resulting distribution of mean AUCs as an empirical null; one-sided empirical P values were computed as the proportion of random gene-set AUCs greater than or equal to the signature AUC. Differences in mean AUC between preprocessing strategies (e.g. whole-blood vs neutrophil-adjusted data, or before vs after ComBat adjustment) and between unimodal and multimodal classifiers were assessed using two-sided Wilcoxon signed-rank tests. Associations between principal components and estimated neutrophil content were assessed using Pearson correlation, and we report correlation coefficients and P values. Group comparisons of insulin-resistance rank gene scores between diagnostic groups (AD vs controls) were performed using two-sided Student’s t-tests. Differential expression analyses were performed using samr, with multiple testing correction false discovery rate (FDR) calculated by permutation, and genes were considered differentially expressed at FDR < 0.05. Gene-set enrichment and dot-plot analyses relied on DAVID through clusterProfiler [39]. For visualisation of co-expression structure, we computed sex-stratified pairwise Pearson correlation matrices between genes in the AD classifier, IR, and B-cell signatures on neutrophil-adjusted expression values (Affymetrix and RNA-seq), restricting to genes present on both platforms and plotting the resulting matrices as annotated heatmaps. For classifier evaluations based on repeated stratified 5-fold cross-validation, model performance was estimated by averaging AUC across 50 repeated 5-fold runs (250 test folds in total). When multiple models were compared, P values from paired tests were adjusted using the Benjamini–Hochberg procedure. Unless otherwise indicated, all tests were two-sided, and a significance threshold of P < 0.05 or FDR < 0.05 was applied.

## Results

### Establishing that the new AD transcriptomic data lacked the bias found in previous generations of data

To assess whether the new transcriptomic dataset is suitable for ML classification studies, we randomly selected ‘gene sets’ and used them to try to distinguish between AD and control samples. If a dataset contains unexpected technical bias, then sets of genes randomly picked can yield a greater classification performance than the theoretical AUC=0.5. However, high-quality transcriptomic data with clearly biological signals between cases and controls will not yield a mean AUC of 0.5, due to the interconnected nature of gene expression. We assessed logistic regression performance using 10,000 randomly sampled gene sets of 75 genes each. First, we compared the distribution of binary classification AUCs for the standard normalised GeneTitan data with that for the ANMerge Illumina data [5] (Supplementary Figure 1, Supplementary Table 1). Our analysis demonstrated that the recent ANMerge Illumina data release is problematic, and more so than previously identified (Figure 2A) [22,24]. In contrast, the new GeneTitan data demonstrated substantially less bias.

**Figure 2.**
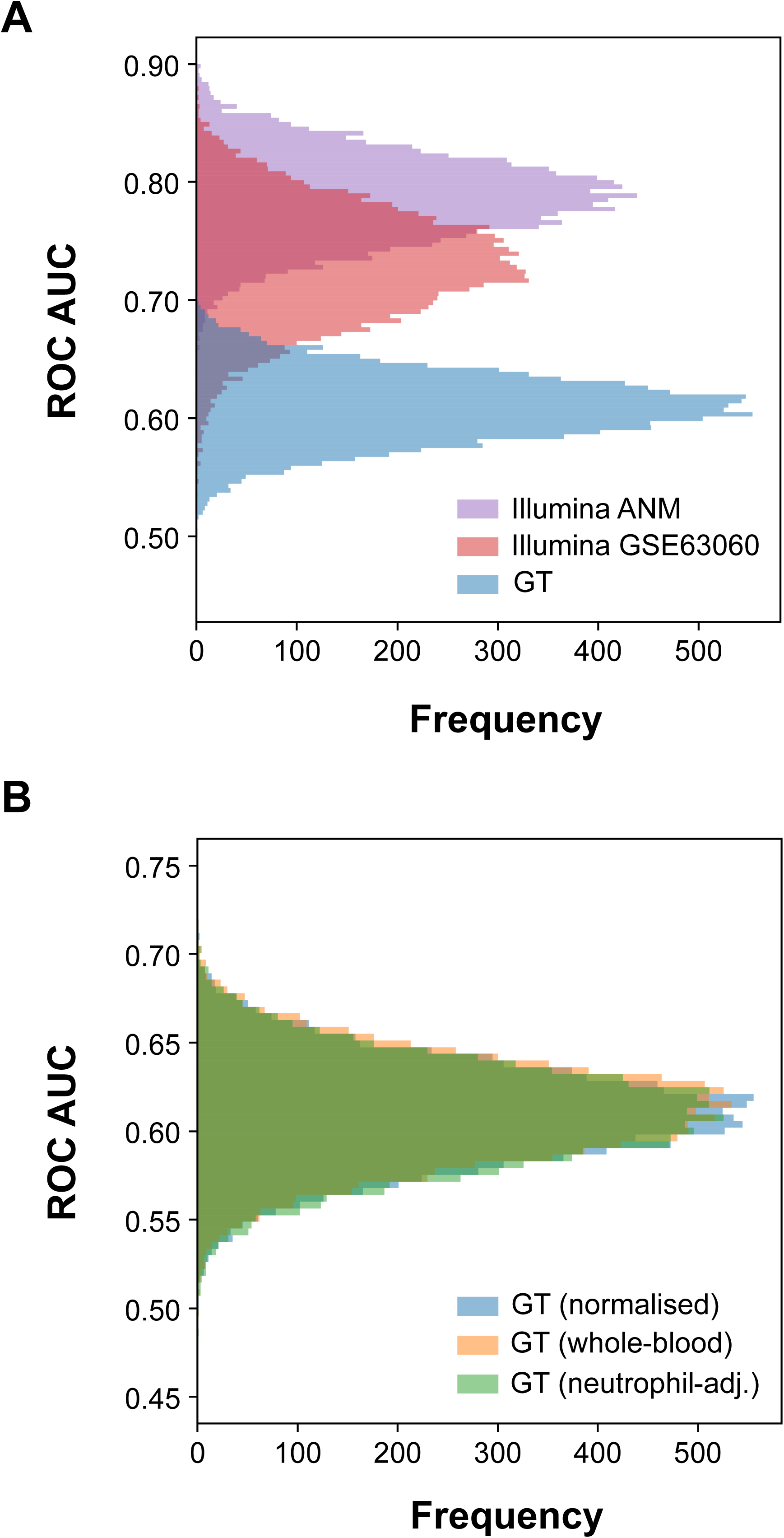
Evaluation of systematic bias in AD and CTL blood transcriptome profiles. Classifiers represented by 75 features were sampled at random from the ANMerge Illumina data source (Illumina ANM, n=162), an updated processing of the AddNeuroMed Illumina data (Illumina GSE63060, n=157) and a new Affymetrix GeneTitan transcriptomic data source (GeneTitan, n=610). Note that the GSE63060 raw data is from the same source data used by ANMerge, but was realigned and annotated to the latest genome in 2024. The third dataset was produced on an Affymetrix GeneTitan (GeneTitan) platform using an HTHGU133Plus PM array. **A**: The distribution of AUCs obtained from 10,000 iterations of sampling at random in the ANMerge reprocessed Illumina ANM data (purple), the original Illumina GSE63060 (red) data (reprocessed – see Methods) and the GeneTitan transcriptomic data (blue). **B**: Investigation of the influence of batch correction for technical variables and whole blood cell composition on the distribution of AUCs obtained over 10,000 iterations of classification using sampling at random; for the standard normalised GeneTitan transcriptomic data (blue), the GeneTitan transcriptomic data adjusted for two technical variables using COMBAT (plate total signal and clinical site (orange) and then this technically adjusted GeneTitan data adjusted for scaled neutrophil counts obtained from absolute immune signal deconvolution (green). Abbreviations: AD, Alzheimer’s disease; CTL, control; AUC, area under the receiver operating characteristic curve.

We examined the impact of adjusting gene expression data for technical variables (total plate signal and clinical centre) and for estimated neutrophil counts (Figure 2B). Stepwise ComBat adjustment for plate signal, clinical centre and then estimated neutrophil counts (Supplementary Figure 2A and 2B) was performed while protecting the primary clinical group label. Adjustment for technical variables led to only a slight increase in the mean classification performance of randomly sampled gene sets (0.609 ± 0.03 to 0.612 ± 0.03 AUC; two-sided Wilcoxon *P* < 0.001). Sequential adjustment for variation in neutrophil counts led to a decrease in random-gene-set performance (0.608±0.03 AUC, Wilcoxon P<0.001). When the primary group labels were not protected during Combat batch correction, we observed a benefit of neutrophil correction on classification (see Discussion). The new GeneTitan data also showed robust technical replicate performance, with better mean global within-replicate pairwise correlation (R=0.954±0.02) than between unrelated samples (R=0.937±0.02).

### Differential gene expression and enrichment analysis

Using the new GeneTitan transcriptomic data, we assessed the impact of cell composition and sex on AD-associated biological pathways (Supplementary Table 2). Neutrophil content [17,35], dominated global transcriptomic variation (Supplementary Figure 2A) e.g. principal component 1 (PC1) (R=0.45, *P*<0.001) and PC2 (R=-0.61, *P*<0.001; See supplementary Figure 2A. On average, females had a higher estimated neutrophil count (*P*<0.001), while mean levels in males did not differ between AD and controls (Supplementary Table 6). When analysing the original Illumina data [13], the authors reported that only basophils were statistically elevated in AD. As basophils represent less than 3% of all white blood cells, this shift should not meaningfully influence global transcriptomic variance. They also observed a 5% numerical increase in neutrophil count (Figure 6A in Lunnon et al. [13]), concordant with our analysis.

Pathway analyses found modest evidence for altered mitochondrial and OXPHOS biology in AD blood, and the strength of this signal varied with the database used and with adjustment for cell composition (Supplementary Tables 7 and 8, Figure 3A and 3B). Using the DAVID_BP_ALL ontology, both female and male whole-blood data showed enrichment for downregulated mitochondrial pathways, including mitochondrial gene expression, proton transmembrane transport and electron transport chain (Figure 3A, Supplementary Table 9). ComBat adjustment for neutrophil fraction substantially reduced the strength of mitochondrial pathway enrichment, with only proton transmembrane transport remaining modestly significant and only in women. Metascape analysis reported reduced mitochondrial ontology enrichment in neutrophil-adjusted data only in women (Figure 3B). Examining the nuclear-encoded OXPHOS genes curated in the MitoCarta database (www.broadinstitute.org/), we observed modest downregulation of individual mitochondrial genes (Supplementary Figure 6A, Supplementary Table 10). In the RNA-seq cohort [26], genes from mitochondrial pathways identified in the array data showed modest changes in expression (Supplementary Figure 7, Supplementary Table 11), and this was similar in both whole-blood and neutrophil-adjusted datasets, with little evidence for sex-specific differences. Previously, Lunnon et al. [11] used qPCR and a ribosomal house-keeping gene to report that mitochondrial DNA (mtDNA) encoded transcripts were *upregulated* in AD. The new GeneTitan data profile six mtDNA genes (where the Illumina data lacks mtDNA probes) and avoids assumptions using ribosomal RNA housekeeping genes. We found that mtDNA RNA expression was unaltered in AD versus control comparisons, irrespective of data processing (Supplementary Figure 6B).

**Figure 3.**
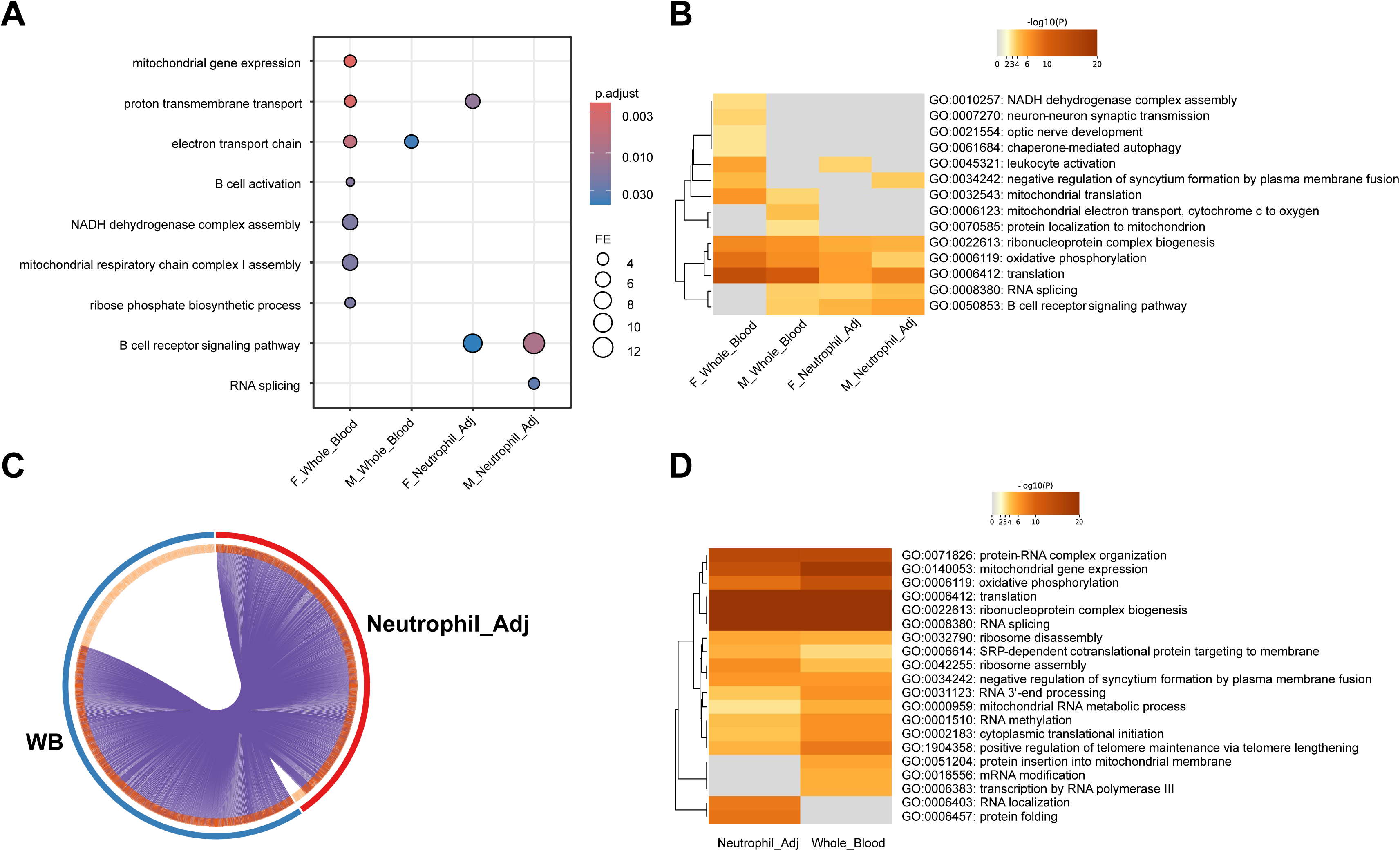
AD-associated mitochondrial and B-cell pathways in whole blood enrichment analyses across sex and neutrophil adjustment. **A:** Dot plot of significant DAVID GOTERM_BP_ALL biological process categories enriched among differentially expressed (DE) genes in female whole blood, male whole blood, female neutrophil-adjusted blood and male neutrophil-adjusted blood (AD vs control); dot size represents fold enrichment and colour indicates adjusted P value (see Supplementary Table 9 for Clusterprofiler results). **B:** Metascape-based Gene Ontology enrichment of DE genes from the same contrasts, showing that both resources highlight pathways related to mitochondrial function, oxidative phosphorylation, ribonucleoprotein complex biogenesis, RNA splicing and B cell receptor signalling. **C:** Summary of the overlap of DE genes between whole blood (WB) and neutrophil-adjusted (Neutrophil_Adj) data, demonstrating that most genes consistently DE in men and women in Neutrophil_Adj are also consistently regulated in WB. **D:** Metascape pathways enriched in the shared set of 2,723 highly consistent genes (less than 10% difference in delta expression between men and women, see Supplementary Table 12), identifying protein translation, RNA processing and mitochondrial processes regardless of data processing. All analyses used the 28,423 transcript DE input list as background (see Supplementary Table 2). Abbreviations: AD, Alzheimer’s disease; CTL, control; Neutrophil_Adj, neutrophil-adjusted; WB, whole blood.

In whole blood, we observed enrichment of B-cell activation pathways only in men (Figure 3A), but following adjustment for neutrophils, B-cell receptor signalling emerged as a feature in both men and women (Supplementary Table 9). These findings were consistent using the Metascape database, which also identified B-cell receptor signalling as more strongly enriched after neutrophil adjustment, in both sexes (Figure 3B). Alterations in the B-cell pathway in AD were confirmed using the Miami RNA-seq cohort, where both B-cell activation and B-cell receptor signalling were enriched in AD (Figure 4). Correlation analyses across the GeneTitan and RNA-seq cohorts indicated that B-cell genes formed a similar co-expression pattern in both sexes across platforms and processing (Supplementary Figures 8 and 9). Overall, these results point to consistent B-cell pathway changes in AD blood across platforms, with some evidence that the strength of enrichment varies by sex.

**Figure 4.**
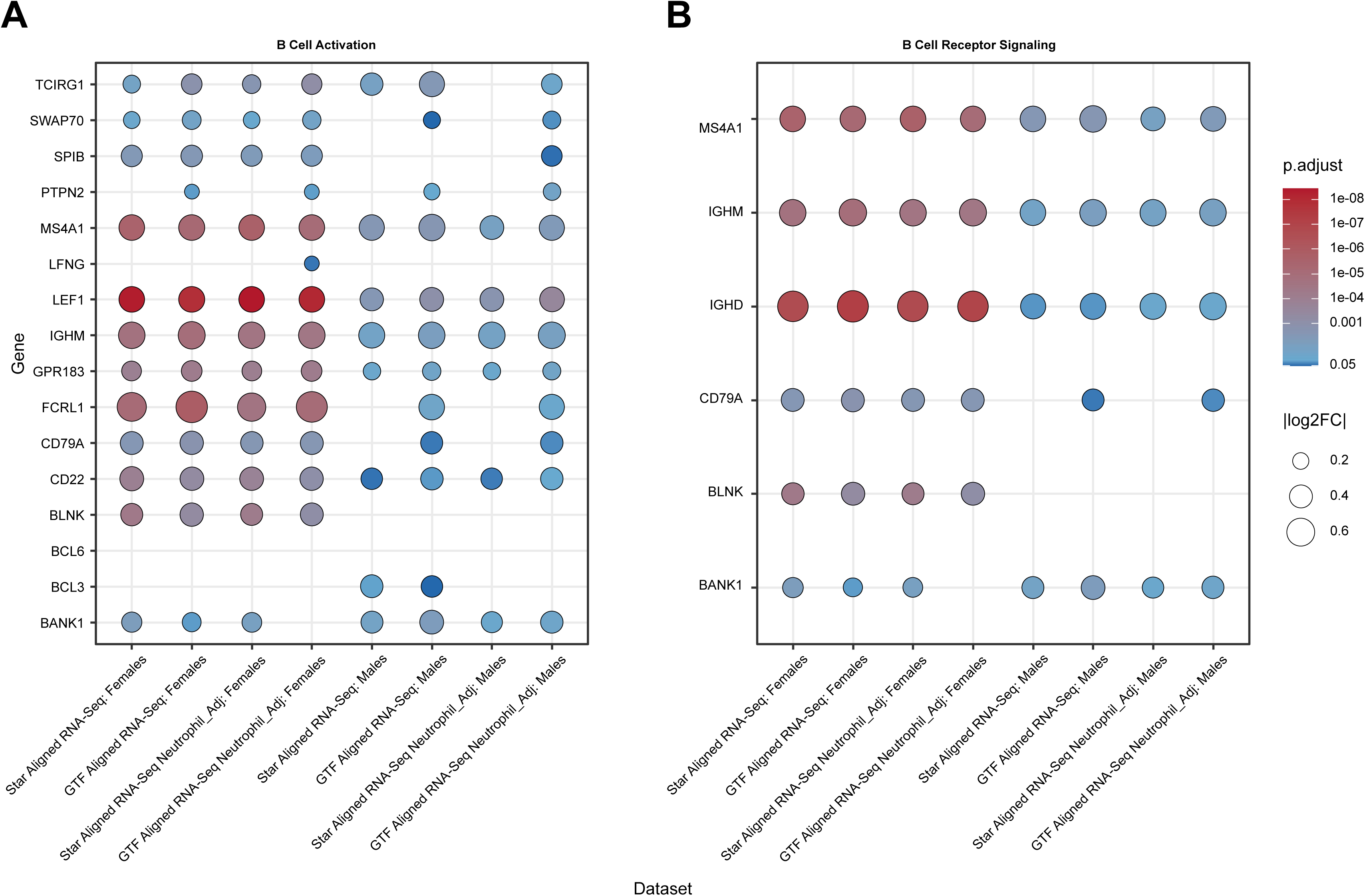
Differential expression of B cell–related genes across AD RNA-seq pipelines by sex. Dot plots show DESeq2 results for genes mapping to two Gene Ontology biological processes: **A:** B cell activation, and **B:** B cell receptor signalling. Rows correspond to individual genes and columns to sex-stratified AD versus control contrasts from four RNA-seq pipelines (STAR-aligned or GTF-aligned, with and without neutrophil adjustment). Dot size encodes the absolute log2 fold change (|log2FC|), and dot color indicates the Benjamini–Hochberg–adjusted P value for each gene in each contrast. Abbreviations: AD, Alzheimer’s disease; DE, differentially expressed; Neutrophil_Adj, neutrophil-adjusted; RNA-seq, RNA sequencing.

To address the limitations of fixed DE thresholds for comparing men and women, we also used hypergeometric analysis (Supplementary Figure 10, Supplementary Table 12) to assess sex differences in AD-associated DE. This analysis revealed strong global consistency for genes downregulated in AD but not upregulated. Genes showing the most consistent down-regulation in both sexes included translation, aspects of mitochondrial biology and RNA splicing (Figure 3C–D). However, most of the concurring differences in downregulated genes were very modest in magnitude. Indeed, Lunnon et al. [13] reported DE of approximately 2,480 genes, but only ∼200 demonstrated >25% difference between groups (Supplementary Table 13), and that is before accounting for the technical bias in that Illumina dataset (Figure 2A). In summary, the pathway features of AD are somewhat consistent in men and women, with apparent sex differences being sensitive to batch correction or methods utilised.

### GeneTitan transcriptomic data for AD classification and multimodal integration

While interpretation of pathway biology is complicated by shifts in whole blood cell composition, our new data demonstrated much lower systematic bias (Figure 2A). We therefore built ML classifiers to distinguish between AD and controls, using logistic regression, support vector machine and random forest models. Using whole blood or neutrophil-adjusted data, supervised feature selection identified a 43-gene whole blood and 44-gene neutrophil-adjusted transcriptomic signature (Supplementary Table 14). Across all three algorithms (Table 4), both whole-blood and neutrophil-adjusted classification models performed similarly. Support vector machine models performed best in both datasets, achieving AUCs of 0.740 (whole-blood) and 0.749 (neutrophil-adjusted). Subjects with paired MRI data were held out for testing, such that the reported AUCs may be more conservative than those obtained under a standard stratified split. Regardless, within-cohort performance estimates of this nature should always be interpreted with caution and confirmed in an independent external cohort (See below).

**Table 4.** AD vs. CTL classification performance of whole blood and neutrophil-adjusted transcriptomics evaluated held-out data.

| **Dataset** | **Model** | **Features** | **AUC** | **AUC. (F)** | **AUC. (M)** | **Sens.** | **Spec.** |
| --- | --- | --- | --- | --- | --- | --- | --- |
| Whole blood | Logistic regression | 43 | 0.736 | 0.738 | 0.757 | 0.580 | 0.679 |
| Neutrophil-adjusted | Logistic regression | 44 | 0.743 | 0.753 | 0.752 | 0.654 | 0.679 |
| Whole blood | Support vector machine | 43 | 0.740 | 0.741 | 0.757 | 0.543 | 0.679 |
| Neutrophil-adjusted | Support vector machine | 44 | 0.749 | 0.760 | 0.754 | 0.630 | 0.691 |
| Whole blood | Random forest | 43 | 0.720 | 0.743 | 0.728 | 0.568 | 0.716 |
| Neutrophil-adjusted | Random forest | 44 | 0.727 | 0.744 | 0.734 | 0.630 | 0.691 |

Blood transcriptome signatures were used to train logistic regression, support vector machine, and random forest models. Models were trained on data for 610 subjects (AD, n = 305; CTL, n = 305) and evaluated on 162 subjects (AD, n = 81; CTL, n = 81). Abbreviations: AUC, AUC, area under the receiver operating characteristic curve; Sens., sensitivity; Spec., specificity; AUC. (F), AUC for female subjects; AUC. (M), AUC for male subjects.

To assess multimodal data integration, we concatenated transcriptomic features with 132 processed MRI features (FreeSurfer 6.0) and compared performance to unimodal models. Model performance was estimated using 50 iterations of 5-fold cross-validation, averaging across 250 test folds. This will be optimistic compared with external validation, which requires new multimodal AD cohort data. Multimodal support vector machine classifiers achieved the best performance (Table 5), with the highest AUC when trained on MRI combined with either whole blood or neutrophil-adjusted data (Supplementary Table 15). Integrating transcriptomics with MRI produced only a modest improvement (whole blood: 0.942 vs. 0.929 AUC, Wilcoxon P<0.001; neutrophil-adjusted: 0.939 vs. 0.929 AUC, Wilcoxon P<0.001). Consistent with sampling-at-random (Figure 2B) and unimodal transcriptomics classifiers (Table 4), within-cohort multimodal performance was robust to sequential adjustment for neutrophil fraction (0.942 vs. 0.939, Wilcoxon *P*<0.001). Overall, the added value of integrating transcriptomic and MRI data for AD classification appears modest in the ANMerge cohort with the caveat that there is no external validation of the MRI contribution.

**Table 5.** AD vs. CTL classification performance of multimodal SVM models integrating blood transcriptomics with MRI data.

| **Multimodal** | **Model** | **Features** | **AUC** | **AUC. (F)** | **AUC. (M)** | **Sens.** | **Spec.** |
| --- | --- | --- | --- | --- | --- | --- | --- |
| No | RNA (Whole blood) | 43 | 0.760 | 0.759 | 0.772 | 0.778 | 0.608 |
| No | RNA (Neutrophil-adjusted) | 44 | 0.748 | 0.738 | 0.770 | 0.751 | 0.615 |
| No | MRI | 132 | 0.929 | 0.914 | 0.958 | 0.810 | 0.884 |
| Yes | RNA + MRI (Whole blood) | 175 | 0.942 | 0.930 | 0.970 | 0.851 | 0.878 |
| Yes | RNA + MRI (Neutrophil-adjusted) | 176 | 0.939 | 0.925 | 0.969 | 0.846 | 0.863 |

Models were evaluated using 50x repeated 5-fold cross-validation on 162 subjects (AD, n = 81; CTL, n = 81). Abbreviations: SVM, support vector machine; AUC, AUC, area under the receiver operating characteristic curve; Sens., sensitivity; Spec., specificity; AUC. (F), AUC for female subjects; AUC. (M), AUC for male subjects.

Genes identified by classification, in both whole-blood and neutrophil-adjusted data, were also enriched in B-cell pathway biology, consistent with our enrichment analysis (Supplementary Tables 9 and 11). We used SHAP [45], to characterise feature-level contributions in the best-performing models. In the neutrophil-adjusted transcriptomics support vector machine model, SHAP values on the independent array test set indicated a broadly distributed pattern of importance, with immune/B cell–associated, metabolic/mitochondrial and chromatin/gene-regulatory transcripts each contributing substantially, consistent with a multifactorial peripheral signature rather than reliance on a small set of dominant genes (Figure 5).

**Figure 5.**
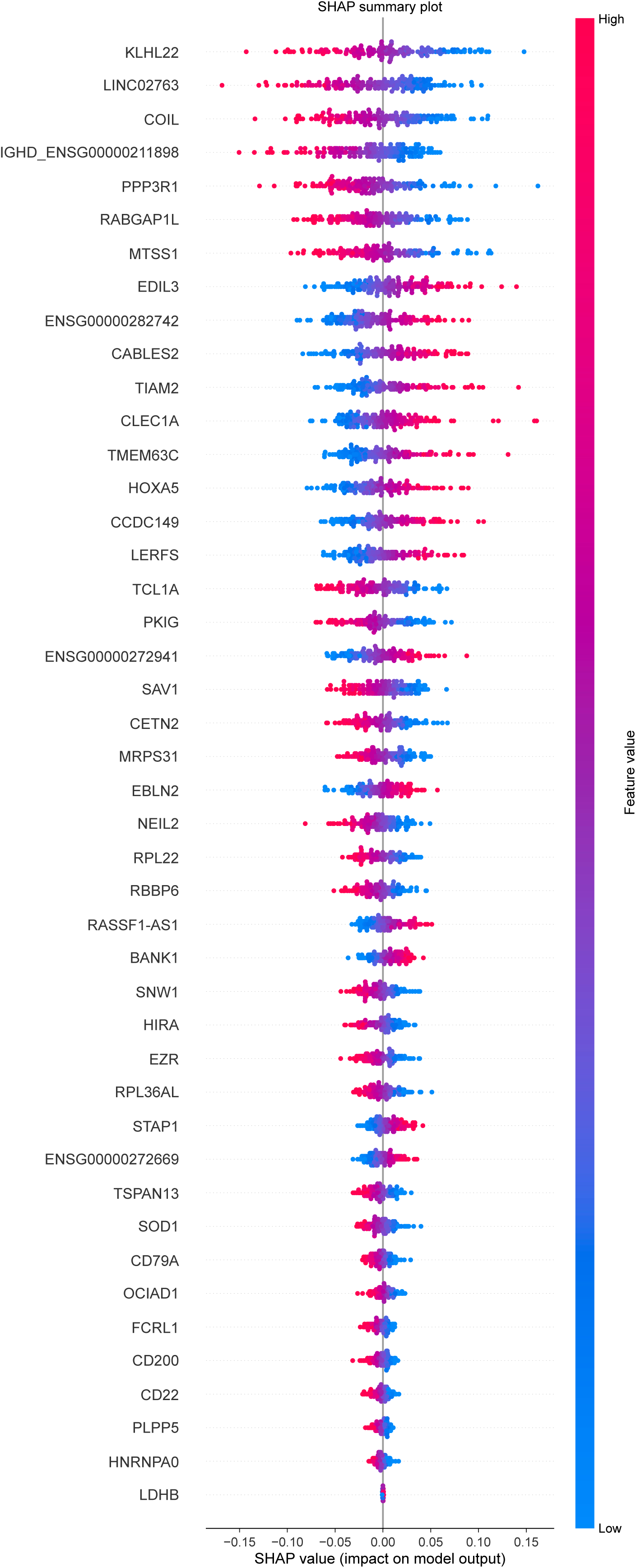
Assessment of the contribution of transcriptomic gene features to model performance. Shapley feature importance values were computed for input features for the best neutrophil-adjusted transcriptomics model (support vector machine) evaluated using the held-out test set (n=162; AD: 81, CTL: 81). Features are ordered by absolute mean Shapley value. Each dot represents a prediction, where a positive Shapley value indicates the feature is positively correlated with the AD class. Each dot’s colour represents the relative feature value. Abbreviations: AD, Alzheimer’s disease; CTL, control.

### External validation of AD blood transcriptome signatures

External validation was carried out using repeated stratified 5-fold cross-validation applied to a class-matched subset of our RNA-seq cohort (Table 6). The neutrophil-corrected signature outperformed the whole-blood signature in both logistic regression and support vector machine models (Wilcoxon *P*<0.001, FDR<0.001), but not in the random forest model (Wilcoxon *P*=0.970). Support vector machines performed best overall, with a mean AUC of 0.780 in neutrophil-adjusted data trained on features selected from the neutrophil-adjusted array data, compared with 0.765 in whole blood (Wilcoxon *P*<0.001). In contrast to the held-out internal validation of the array cohort, where no sex differences were observed, performance in the external RNA-seq cohort was consistently better in women (Supplementary Figure 11). Results were similar using genomic GTF-aligned and array GTF-aligned data (Supplementary Table 16). However, as noted, when we applied a hypothesis-generated external signature [51] or a data-driven internal model [24], bias in the original data can inflate the estimated performance. Sampling-at-random (Figure 1E, Supplementary Figure 3A) using the array GTF-aligned, neutrophil-adjusted RNA-seq data, produced a mean AUC across random gene sets of 0.729±0.024 (Supplementary Figure 12). However, only 0.22% of sampling-at-random sets achieved an AUC that matched our externally derived model (≥0.788, *P*=0.002), and that is without removing our AD-associated genes. In whole-blood RNA-seq data, the mean random AUC was 0.730, closer to our AD classifier signature (AUC=0.767, *P*=0.06), indicating that neutrophils inflate AUC estimates in the RNA-seq cohort. Overall, these results emphasise that blood cell biases can induce global structure that ML exploits. [33,52,53].

**Table 6.** AD vs. CTL classification performance of GeneTitan array transcriptomic signatures evaluated on independent RNA-seq.

| **Dataset** | **Model** | **Features** | **AUC** | **AUC. (F)** | **AUC. (M)** | **Sens.** | **Spec.** |
| --- | --- | --- | --- | --- | --- | --- | --- |
| Whole blood | Logistic regression | 39 | 0.757 | 0.771 | 0.730 | 0.679 | 0.714 |
| Neutrophil-adjusted | Logistic regression | 39 | 0.773 | 0.791 | 0.740 | 0.718 | 0.729 |
| Whole blood | Support vector machine | 39 | 0.768 | 0.775 | 0.749 | 0.665 | 0.744 |
| Neutrophil-adjusted | Support vector machine | 39 | 0.788 | 0.802 | 0.760 | 0.735 | 0.737 |
| Whole blood | Random forest | 39 | 0.742 | 0.756 | 0.716 | 0.680 | 0.695 |
| Neutrophil-adjusted | Random forest | 39 | 0.741 | 0.769 | 0.687 | 0.684 | 0.681 |

Whole blood and neutrophil-adjusted signatures were evaluated on logistic regression, support vector machine, and random forest models using 10 iterations of 5-fold stratified cross-validation. Models were evaluated on data for 372 subjects (AD, n = 186; CTL, n = 186). Abbreviations: AUC, AUC, area under the receiver operating characteristic curve; Sens., sensitivity; Spec., specificity; AUC. (F), AUC for female subjects; AUC. (M), AUC for male subjects.

When finalising our work, we noticed a recent meta-analysis that used Illumina AddNeuroMed and ADNI data along with a large, unpublished AD data set [27,54]. Each data set was produced on very distinct platforms, aligned to genome builds from across decades, and no attempt was made to harmonise the validity of the gene annotations. Annotation changes due to genomic realignment remove candidate biomarkers, e.g. almost a third of our hypothesis-driven signatures [51] are no longer transferable onto the new data. Reliance on matching using gene symbols is not an acceptable solution because, after realignment, probes may target multiple genes. One large and novel data set (N=453 AD and Control samples) was produced using the Illumina platform [54]. The Nachun et al. [54] data set was released as a pre-print in 2019 and has not yet been published. Applying the sampling-at-random method described above to the batch-corrected data, we found a mean AUC of 0.53 (Supplementary Figure 13A), indicating the performance of most gene-sets was close to theoretical random (even though some should include AD-regulated genes). We noted that Nachun et al. [54] used a variety of data transformations. Likewise, Hou et al. [27] who used the Nachun et al. data (GSE140829) as a validation cohort in a meta-analysis, applied a variety of supervised and unsupervised batch corrections. When we examined the top five AD differentially expressed genes from the Hou et al. [27] meta-analysis, they were invariant in our new data (Supplementary Figure 13B), and when replotted using the Nachun et al. normalised adjusted data (GSE140829), no difference between AD and control groups was observed (Supplementary Figure 13C). We believe these observations are consistent with the signal between cases and controls, including any AD-related signal, being removed during preprocessing.

### IR transcriptome signature reveals systemic differences between AD and controls

To assess whether IR-status assessed using a blood RNA biomarker differed between AD cases and controls, we used an independent IR transcriptomic signature derived from our metabolic physiology cohorts (Supplementary Figure 5A–C). A score based on these 62 genes (Supplementary Table 4) segregated samples from the two log2HOMA2-IR groups in the SPD GeneTitan data (Supplementary Figure 5F) and stratified the S2 U133PM samples across the log2HOMA2-IR range (Supplementary Figure 5G; Spearman ρ=0.438, *P*<0.001). As expected, the correlation in the model development cohort remained robust (Supplementary Figure 5E; Spearman ρ=0.659, *P*<0.001). Genes in the core 62-gene set included several implicated previously in the biology of IR (see Discussion). Fifty-one genes from the IR signature were present in the AD GeneTitan cohort data, and IR rank gene scores were computed in the neutrophil-adjusted dataset. Surprisingly, control samples from AddNeuroMed had higher IR gene scores than AD cases (Figure 6), and this pattern was consistent in women (t-test *P*=0.0274) and men (t-test *P*=0.0288). This indicates that controls were, on average, metabolically less healthy, and on reflection, we believe this may reflect the recruitment of family carers of AD subjects and the known metabolic impact of dementia caregiving [55]. Correlation analyses across the GeneTitan and RNA-seq cohorts further indicated that IR classifier genes showed similar co-expression patterns in men and women across platforms, but limited evidence that the IR signature genes correlated with the AD-linked B-cell pathway genes (Supplementary Figures 7 and 8) or with the AD classifier model genes (Supplementary Figures 14 and 15). This indicates that the IR signatures capture largely distinct aspects of the blood transcriptome from AD.

**Figure 6.**
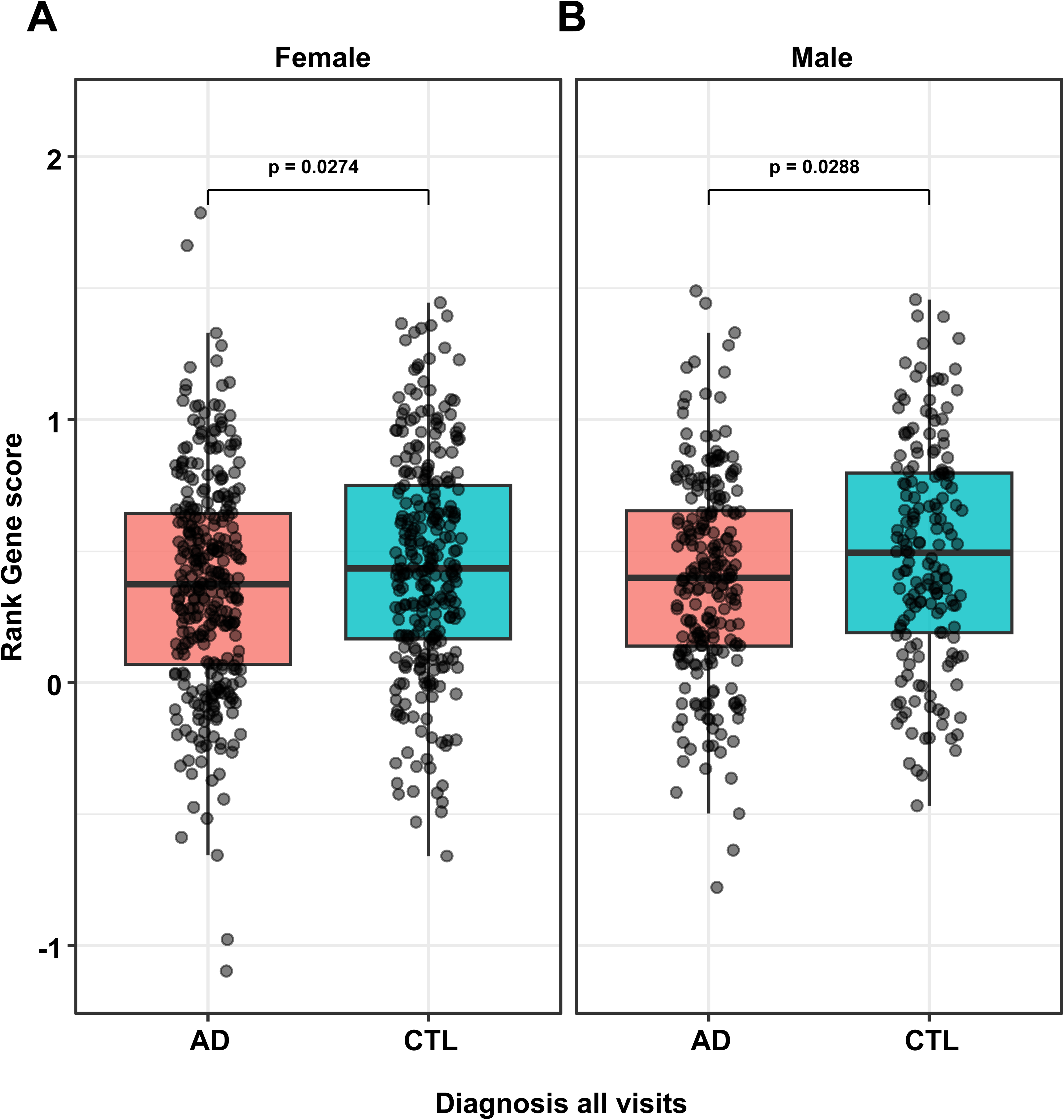
Sex-stratified distribution of insulin resistance rank gene scores in the AD GeneTitan cohort. IR rank gene scores were computed in the neutrophil-adjusted GeneTitan data using the subset of 51 genes from the 62-gene cross-cohort IR transcriptomic signature that were detected on the array, with higher values indicating a more IR-like blood transcriptomic profile. A: Male subjects (n = 385, n(AD) = 222, n(CTL) = 163). B: Female subjects (n = 579, n(AD) = 306, n(CTL) = 273). P values from two-sample t tests are shown above each comparison and suggest modestly higher IR rank gene scores in controls compared to AD, consistent in women and men. Abbreviations: AD, Alzheimer’s disease; CTL, control; IR, insulin resistance.

## Discussion

We present a new, large and robust AD transcriptomic resource. The original blood transcriptomics from overlapping samples have technical limitations [22,24], and these issues remain pronounced even after recent processing (Figure 2A). The new GeneTitan data can be realigned to the genome in future years, to remain relevant. The equivalent ADNI data cannot be updated [32] because the original probe-level data are no longer available. Our new resource enabled the first robust evaluation of AD-linked sexual dimorphism, and we find that the influence of white blood cell content and sex on the molecular pathways associated with AD is complex. Our findings do not support the original claims [11] that mtDNA-encoded transcripts are up-regulated in AD or that there is a lack of influence of blood cell composition on the transcriptome. The new data were collected predominantly in white participants [56], and recruitment of subjects from underrepresented populations remains a challenge in AD research [57]. However, we find that our blood AD classification signature retains some discriminative performance in an independent RNA-seq cohort (distinct technology and ethnicities), and that key biological themes from differential expression analysis were reproducible. We also find that blood cell composition and cohort-specific structure will bias machine-learning models if not carefully checked.

Evaluation of the transcriptomic classifier genes, in the neutrophil-adjusted GeneTitan data set, using Shapley value–based feature attribution, on the held-out test set, indicated as expected that the signature draws on multiple biological processes rather than a single dominant pathway (Figure 5). Among higher-ranking transcripts, we observe enrichment for processes involved in synaptic plasticity, calcium-dependent signalling, and neurovascular regulation. Notably, *PPP3R1*, which encodes the regulatory subunit of the phosphatase calcineurin, was found to be upregulated in the AD state. Calcineurin (also a classic T-Cell signalling molecule) hyperactivation is a driver of Aβ-induced synaptic dysfunction, tau hyperphosphorylation, and neuroinflammation [58]. Furthermore, genes critical for cytoskeletal dynamics and structural synaptic plasticity were present. Specifically, *TIAM2*, a regulator of Rac1-mediated dendritic spine morphogenesis, was downregulated in the disease cohort, providing a potential molecular marker for the progressive synaptic loss characteristic of AD pathology [59]. Upregulation of *EDIL3* suggests ongoing vascular or endothelial alterations, aligning with the growing recognition of neurovascular unit compromise and altered angiogenesis in the progression of the disease [60]. Given the instability of single-gene ranking-based feature selection in high-dimensional transcriptomic data and the influence of cohort-specific variance, we interpret these genes as illustrative examples of broader mechanistic themes rather than definitive blood biomarkers, and independent validation will be needed to confirm their specific roles.

There remains a need for higher performing AD diagnostic biomarkers derived from samples obtained using minimally invasive methods, and further work with the new resources presented herein will assist with that process. Robust biomarkers, such as those using CSF, remain less practical, as only 40% of individuals are willing to undertake a lumbar puncture, compared with 75% willing to have an MRI or PET scan, or 81% for blood tests, with women less willing to have a lumbar puncture than men (37% of women vs. 48% of men) [61]. Recent studies show that plasma p-tau^217^ demonstrates diagnostic accuracy equivalent to CSF p-tau^217^, indicating that plasma-based markers can act as a viable alternative to CSF markers in detecting AD pathology [8]. Elevated plasma p-tau^181^ has also been associated with poorer neuropsychological test performance, based on retrospective analysis of subjects from the ADNI dataset [7]. These biomarkers do not provide new information on disease pathology, and so the application of genome-level profiling of human tissue and blood remains an important objective [27]. Our analysis found consistent evidence that immune cell composition influences global variance in whole-blood transcriptomic data and can obscure disease-specific signal if not modelled explicitly [62]. Applying batch adjustment without protecting for primary group variables modestly decreases within-cohort performance, highlighting that statistical adjustment may minimise disease-related signal. In line with this, when we examined a recent unpublished AD whole-blood transcriptomic cohort, we noted that the processed data showed essentially no evidence of candidate AD gene differential expression, consistent with excessive batch correction or confounded design removing disease-related signal.

We observed greater classification performance in women in the RNA-seq external cohort, despite sex-stratified enrichment analyses indicating that modulated pathways, such as B-cell signalling, were similar in both sexes. Additionally, neutrophil adjustment improved independent performance more in women than in men, but the mechanisms underlying this sex difference remain unclear. An important observation in our study is the influence of cell proportions on pathway biology, particularly neutrophils, which typically comprise 40–60% of peripheral blood leukocytes in a whole blood sample [63]. While adjustment for cell composition is important for ML classification, it complicates the interpretation of AD-associated pathway biology. Neutrophil-to-lymphocyte ratio (NLR) is a crude marker of inflammatory status in cancer, cardiovascular and other inflammatory diseases [64,65]. Chronic elevation of pro-inflammatory cytokines is associated with greater cognitive decline in AD [66], and elevated neutrophil levels have been reported in neurodegenerative diseases such as PD and in chronic conditions prevalent in AD populations, such as type 2 diabetes [53,67]. These observations indicate that altered whole-blood neutrophil content cannot be disease-specific for AD, and thus a barrier for developing a specific diagnostic. [16]

In AD, NLR was initially reported to correlate with amyloid burden in the AIBL cohort, but this association did not remain after adjustment for age, sex and APOE ε4 allele status [16]. Despite the lack of AD *specificity*, neutrophils may still be involved in AD pathogenesis. Recent evidence suggests that neutrophils infiltrate the brain in AD, migrate toward amyloid plaques and promote blood–brain barrier disruption, capillary flow stalling and neuroinflammation, in part through neutrophil extracellular trap (NET) formation [68,69]. Clinical studies also indicate neutrophil hyperactivation in AD, with increased reactive oxygen species production, elevated intravascular NETs and altered neutrophil phenotypes that correlate with disease progression [70]. NETs have been directly observed in cortical blood vessels and brain parenchyma of AD patients, supporting a mechanistic role in vascular and neural damage [68]. Thus, our statistical concerns about neutrophil proportions as confounders in ML models do not preclude a mechanistic role for neutrophils in AD pathogenesis. By explicitly accounting for white blood cell composition, we highlight how such factors can impact the disease specificity of blood-based AD diagnostics, without directly judging their role in AD pathophysiology.

We present novel evidence of immune and inflammatory pathway modulation in both women and men with AD. The independent RNA-seq revealed a core set of consistently upregulated immune genes across multiple processing pipelines (Supplementary Table 11), including genes directly involved in B-cell receptor signalling (CD79A, BLNK) and regulation of B-cell activation (BANK1). [71,72]. In triple transgenic mouse models of AD, B-cells are implicated in reducing Aβ plaques and accelerating neuroinflammation, with effects driven by specific B-cell subtypes [73]. B-cell depletion also worsened both spatial learning and memory deficits in AD mice and was associated with increased Aβ burden [74]. Conversely, B-cell-deficient mice showed elevated IgG near Aβ plaques and increased microglial activation, with further B-cell depletion reducing amyloid deposition and improving cognitive function [75]. These findings suggest that B cells may have context-dependent, potentially neuroprotective roles in early AD pathology, including a relationship with production of the anti-inflammatory cytokine interleukin-35 [74]. Whether analysis of whole blood can reveal sufficiently specific molecular mechanisms underlying B-cell involvement in AD remains to be determined. Given the nearly doubled lifetime risk of AD in women compared with men (approximately 1 in 5 versus 1 in 10 at age 45) [76], we explicitly examined the sex-stratified behaviour of our blood signatures. However, after adjusting for immune composition and technical covariates, we did not identify robust, cross-platform sex-specific modulation of the identified B-cell or classifier gene sets in blood. Co-expression structures were highly similar between sexes, and sex-stratified differential expression patterns for the classifier genes showed broadly concordant AD–control effects.

Our cross-cohort analysis identified a 62-gene IR transcriptomic signature. Application of this signature to the AD blood cohort revealed higher IR-like scores in controls than in AD cases, in both women and men. Research cohorts studying AD attempt to control for comorbidities during recruitment, but this will not be the case when a new clinical diagnostic is deployed in the general population. In addition, strategies [28] such as using caregiving spouses as controls [55] may introduce systematic metabolic differences between groups, potentially contributing to the higher IR-related signatures we observed in some control groups in this study. This pattern likely reflects characteristics of the control population [55] rather than a protective effect of IR in AD. The cohort consists of elderly individuals, and AD is typically associated with increased mortality, with poor metabolic health and diabetes further increasing risk [77]. Given the recruitment of AD and control groups with similar age distributions (Table 4), AD cases may be somewhat enriched for individuals who have survived AD so far. Combined, survivorship and recruitment biases could deplete IR individuals among AD cases, despite established epidemiological evidence linking IR and type 2 diabetes to elevated risk of AD and dementia [19]. Notably, the IR and AD classification signatures do not show strong cross-correlation (Figure S12 and S13), meaning both could be utilised for drug trial cohort enrichment.

The IR transcriptomic signature itself captured well-characterised metabolic pathways with direct and well-characterised roles in metabolism. *KCNQ1* variants are repeatedly associated with type 2 diabetes and impaired insulin secretion [78], and functional studies demonstrate that *KCNQ1* influences insulin signalling through the IRS-2/PI3K/Akt pathway, affecting insulin sensitivity and therapeutic response in hepatocyte models [79]. Similarly, loss or dysfunction of *LDLR* leads to dyslipidemia and hepatic IR in animal models, where *LDLR*-deficient mice and rats develop glucose intolerance and IR when exposed to Western-type or high-fat diets [80–82]. Other genes in the signature link to insulin physiology and downstream metabolic processes that are central to insulin action. *EXOC6*, a component of the exocyst complex, couples insulin signalling to vesicle trafficking by interacting with Rab10-GTP to regulate GLUT4 translocation and insulin-stimulated glucose uptake in adipose tissue and muscle [83]. Disruption of this pathway provides a clear mechanistic route to IR. In addition, *IFNAR2* and *GATA2* contribute more indirectly through immune and vascular pathways that influence islet inflammation, β-cell function, and metabolic homeostasis, further reinforcing the biological relevance of the signature genes to IR–associated phenotypes [80,84,85].

Multimodal learning can enhance performance in disease classification and progression prediction tasks [86,87]. Using concatenation, a simple fusion approach [87], we found that blood RNA profiles produced only modest improvement over unimodal models trained on MRI features (Supplementary Table 15). The relationship between structural MRI features and Illumina blood transcriptomics in AddNeuroMed was previously examined using linear regression between PCA-based module eigengenes and MRI measures, reporting a positive association between a mitochondrial (OXPHOS) module and hippocampal volume. Whether this association remains after accounting for blood cell composition and technical confounding is unknown. Maddalena et al. [88] also used ANMerge MRI and transcriptomics, reporting that concatenation improved classification performance. In contrast, our analysis using bias-corrected data shows only modest improvement, suggesting that previously reported gains may be influenced confounding transcriptomic data (Figure 2A and 2B). Likewise, without external data, it is unclear what the true performance of MRI methods will be.

## Conclusions

We provide an expanded and technically more robust blood transcriptomic resource for the ANMerge database, suitable for machine learning applications and large enough to interrogate sex-specific molecular features of AD. Using this new data, we show that a microarray-derived data generalises to an independent RNA-seq cohort. We find immune cell proportions are a notable source of group-specific variance in blood-based transcriptomic classifiers. Our sex-stratified analyses reveal consistent B-cell–related pathway modulation in women and men with AD, which was more robust after adjusting for immune composition. We also demonstrate consistent female advantage in classification performance in independent validation, in contrast to DE methods, suggesting that further analytical approaches are needed to fully explore sex differences. Further, applying a cross-cohort insulin-resistance transcriptomic signature to the AD blood cohort revealed higher IR-like scores in controls, which should be integrated into any future modelling. Together, these findings emphasise the need to model cell composition and sex explicitly in machine-learning studies and provide a resource and analytic framework that can be extended to more diverse populations, age-matched metabolic cohorts, and larger multimodal datasets.

## Supporting information

Input data filenames and sources for analyses

## Abbreviations

Aβ: Amyloid β
AD: Alzheimer’s disease
AUC: Area under the receiver operating characteristic curve
CTL: Control
GTF: Gene transfer format
HOMA2-IR: Homeostatic Model Assessment 2 of Insulin Resistance
IR: Insulin resistance
MP: META-PREDICT
MRI: Magnetic resonance imaging
mRMR: Minimum redundancy–maximum relevance
OXPHOS: Oxidative phosphorylation
S2: STRRIDE-AT/RT
SHAP: SHapley Additive exPlanations
SPD: STRRIDE-PD

## Declarations

### Ethics approval and consent to participate

Blood samples for the RNA analyses were obtained from individuals taking part in the biomarker studies (Coordinated by Dr Angela Hodges; The AddNeuroMed study and the Maudsley Biomedical Research Centre Dementia Case Register) as previously reported [11]. The clinical study obtained informed consent according to the Declaration of Helsinki (1991) and ethical approval at each of the six clinical centres (London, Kuopio, Lodz, Perugia, Thessaloniki, and Toulouse). For the independent RNA-seq whole-blood transcriptomic study described by Griswold et al. [26], all participants, or their legally authorised representatives, gave informed consent, and procedures were approved by the University of Miami Institutional Review Board. The multi-centre META-PREDICT cohort was funded by a Seventh Framework Programme (FP7) grant and approved by local ethics committees at each centre (the University of Nottingham Medical School Ethics Committee, the Regional Ethical Review Board Stockholm, the ethics committee of the municipality of Copenhagen and Frederiksberg in Denmark, the Comite Etico de Investigacion Humana de the Universidad de Las Palmas de Gran Canaria, and the Loughborough University Ethics Approvals (Human Participants) Sub-Committee). The STRRIDE AT-RT study protocol was approved by the Institutional Review Boards at Duke University and East Carolina University, and the STRRIDE-PD study protocol was approved by the Institutional Review Board at Duke University.

### Consent for publication

Not applicable. This manuscript does not contain any individual person’s data in any form (including individual details, images or videos) that would require consent for publication.

### Availability of data and materials

The raw gene expression data reported in this paper are available at E-MTAB-15140, along with the processed data files and novel CDFs. The probes for the raw data can be realigned to the current genome and transcriptome each year to remain current. Code for the various informatics analyses can be readily obtained by contacting the authors or via https://github.com/Nasim-MI/Affy-ANMerge-ML. The version of the realigned raw count RNA-seq data used in the present study, together with the associated processed matrices and phenotype data for the Miami cohort, can be accessed via Zenodo (https://doi.org/10.5281/zenodo.20269433).

### Author’s contributions

The study aims and objectives were conceptualised by JAT, GS, NMI, AJG, MM and CHV. Bioinformatics analysis was carried out by NMI, JAS, CK, TG and JAT. Data collection and analysis were done by HC, BEP, RJB, AJG, WEK and colleagues within the AddNeuroMed consortium. The manuscript was drafted by NMI, MM, CV and JAT. Revision of the manuscript was carried out by NMI, MM, HC, PJA, CW, JPC, WEK, KS, AJG, CHV, GS and JAT.

## Acknowledgements

We would like to thank Dr Angela Hodges (King’s College London) and Dr Simon Lovestone for their access to the AD biobank RNA samples, and to all the AddNeuroMed investigators for the blood samples used in this study. We thank Dr Sanjana Sood for helping to prepare the samples for profiling. We thank the former board of Affymetrix Inc, especially Dr Muriel Chapoutot. We acknowledge the use of Queen Mary’s Apocrita HPC facility, supported by QMUL Research-IT. Additional computational resources were provided by Augur Precision Medicine LTD.

## References

1. Long S, Benoist C, Weidner W. World Alzheimer Report 2023: Reducing Dementia Risk: Never too early, never too late [Internet]. Alzheimer’s Disease International; 2023 Dec. https://www.alzint.org/u/World-Alzheimer-Report-2023.pdf. Accessed 11 Dec 2024

2. Cipriani G, Danti S, Picchi L, Nuti A, Fiorino M Di. Daily functioning and dementia. Dement Neuropsychol [Internet]. Academia Brasileira de Neurologia; 2020;14:93–102. 10.1590/1980-57642020dn14-020001

3. Sperling RA, Aisen PS, Beckett LA, Bennett DA, Craft S, Fagan AM, et al. Toward defining the preclinical stages of Alzheimer’s disease: Recommendations from the National Institute on Aging-Alzheimer’s Association workgroups on diagnostic guidelines for Alzheimer’s disease. Alzheimer’s & Dementia [Internet]. 2011;7:280–92. 10.1016/j.jalz.2011.03.003

4. Cummings JL, Morstorf T, Zhong K. Alzheimer’s disease drug-development pipeline: Few candidates, frequent failures. Alzheimers Res Ther. 2014;6:1–7. 10.1186/alzrt269

5. Birkenbihl C, Westwood S, Shi L, Nevado-Holgado A, Westman E, Lovestone S, et al. ANMerge: A Comprehensive and Accessible Alzheimer’s Disease Patient-Level Dataset. Journal of Alzheimer’s Disease [Internet]. IOS Press; 2021;79:423–31. 10.3233/JAD-200948

6. Petersen RC, Aisen PS, Beckett LA, Donohue MC, Gamst AC, Harvey DJ, et al. Alzheimer’s Disease Neuroimaging Initiative (ADNI). Neurology [Internet]. American Academy of Neurology; 2010;74:201–9. 10.1212/WNL.0b013e3181cb3e25

7. Bolton CJ, Steinbach M, Khan OA, Liu D, O’Malley J, Dumitrescu L, et al. Clinical and demographic factors modify the association between plasma phosphorylated tau-181 and cognition. Alzheimer’s & Dementia: Diagnosis, Assessment & Disease Monitoring [Internet]. John Wiley & Sons, Ltd; 2024 [cited 2025 Jan 30];16:e70047. 10.1002/dad2.70047

8. Khalafi M, Dartora WJ, McIntire LBJ, Butler TA, Wartchow KM, Hojjati SH, et al. Diagnostic accuracy of phosphorylated tau217 in detecting Alzheimer’s disease pathology among cognitively impaired and unimpaired: A systematic review and meta-analysis. Alzheimer’s & Dementia [Internet]. John Wiley & Sons, Ltd; 2025 [cited 2025 Jan 30];21. 10.1002/alz.14458

9. Phillips JM, Dumitrescu LC, Archer DB, Regelson AN, Mukherjee S, Lee ML, et al. Novel modelling approaches to elucidate the genetic architecture of resilience to Alzheimer’s disease. Brain [Internet]. Brain; 2025 [cited 2025 Jul 23]; 10.1093/BRAIN/AWAF106

10. Peterson A, Sathe A, Zaras D, Yang Y, Durant A, Deters KD, et al. Sex and APOE ε4 allele differences in longitudinal white matter microstructure in multiple cohorts of aging and Alzheimer’s disease. Alzheimer’s & Dementia [Internet]. John Wiley & Sons, Ltd; 2025 [cited 2026 Apr 27];21:e14343. 10.1002/ALZ.14343

11. Lunnon K, Keohane A, Pidsley R, Newhouse S, Riddoch-Contreras J, Thubron EB, et al. Mitochondrial genes are altered in blood early in Alzheimer’s disease. Neurobiol Aging [Internet]. Neurobiol Aging; 2017 [cited 2025 Mar 12];53:36–47. 10.1016/J.NEUROBIOLAGING.2016.12.029

12. Libby JB, Seto M, Khan OA, Liu D, Petyuk V, Oliver NC, et al. Whole blood transcript and protein abundance of the vascular endothelial growth factor family relate to cognitive performance. Neurobiol Aging [Internet]. Elsevier Inc.; 2023 [cited 2025 Jul 23];124:11–7. 10.1016/j.neurobiolaging.2023.01.002

13. Lunnon K, Ibrahim Z, Proitsi P, Lourdusamy A, Newhouse S, Sattlecker M, et al. Mitochondrial Dysfunction and Immune Activation are Detectable in Early Alzheimer’s Disease Blood. Journal of Alzheimer’s Disease [Internet]. SAGE PublicationsSage UK: London, England; 2012 [cited 2025 Jan 30];30:685–710. 10.3233/JAD-2012-111592

14. Heppner FL, Ransohoff RM, Becher B. Immune attack: the role of inflammation in Alzheimer disease. Nat Rev Neurosci [Internet]. 2015;16:358–72. 10.1038/nrn3880

15. Luo J, Thomassen JQ, Nordestgaard BG, Tybjærg-Hansen A, Frikke-Schmidt R. Blood Leukocyte Counts in Alzheimer Disease. JAMA Netw Open [Internet]. 2022;5:e2235648. 10.1001/jamanetworkopen.2022.35648

16. Rembach A, Watt AD, Wilson WJ, Rainey-Smith S, Ellis KA, Rowe CC, et al. An increased neutrophil–lymphocyte ratio in Alzheimer’s disease is a function of age and is weakly correlated with neocortical amyloid accumulation. J Neuroimmunol [Internet]. Elsevier; 2014 [cited 2025 Feb 4];273:65–71. 10.1016/j.jneuroim.2014.05.005

17. Nath M, Romaine SPR, Koekemoer A, Hamby S, Webb TR, Nelson CP, et al. Whole blood transcriptomic profiling identifies molecular pathways related to cardiovascular mortality in heart failure. Eur J Heart Fail [Internet]. John Wiley and Sons Ltd; 2022 [cited 2025 Feb 25];24:1009–19. 10.1002/ejhf.2540

18. Timmons JA, Atherton PJ, Larsson O, Sood S, Blokhin IO, Brogan RJ, et al. A coding and non-coding transcriptomic perspective on the genomics of human metabolic disease. Nucleic Acids Res. Oxford University Press; 2018;46:7772–92. 10.1093/nar/gky570

19. Cao F, Yang F, Li J, Guo W, Zhang C, Gao F, et al. The relationship between diabetes and the dementia risk: a meta-analysis. Diabetol Metab Syndr [Internet]. BioMed Central Ltd; 2024 [cited 2026 Mar 17];16:101. 10.1186/s13098-024-01346-4

20. Arnold SE, Arvanitakis Z, Macauley-Rambach SL, Koenig AM, Wang HY, Ahima RS, et al. Brain insulin resistance in type 2 diabetes and Alzheimer disease: concepts and conundrums. Nature Reviews Neurology 2018 14:3 [Internet]. Nature Publishing Group; 2018 [cited 2026 Mar 17];14:168–81. 10.1038/nrneurol.2017.185

21. Cousins KAQ, Boyle R, Morse C, Verma A, Brown CA, O’Brien KS, et al. Electronic Health Records to Test Multimorbidity Influences to Plasma Biomarker Interpretation for Alzheimer’s Disease. Ann Neurol [Internet]. Ann Neurol; 2026 [cited 2026 May 22];99. 10.1002/ANA.78114

22. Voyle N, Keohane A, Newhouse S, Lunnon K, Johnston C, Soininen H, et al. A Pathway Based Classification Method for Analyzing Gene Expression for Alzheimer’s Disease Diagnosis. Arendash G, editor. Journal of Alzheimer’s Disease [Internet]. 2015;49:659–69. 10.3233/JAD-150440

23. Lunnon K, Sattlecker M, Furney SJ, Coppola G, Simmons A, Proitsi P, et al. A Blood Gene Expression Marker of Early Alzheimer’s Disease. Journal of Alzheimer’s Disease [Internet]. IOS Press; 2013 [cited 2025 Feb 25];33:737–53. 10.3233/JAD-2012-121363

24. Timmons JA, Gallagher IJ, Sood S, Phillips B, Crossland H, Howard R, et al. A statistical and biological response to an informatics appraisal of healthy aging gene signatures. Genome Biol. BioMed Central Ltd.; 2019;20. 10.1186/s13059-019-1734-z

25. Stokes T, Cen HH, Kapranov P, Gallagher IJ, Pitsillides AA, Volmar C, et al. Transcriptomics for Clinical and Experimental Biology Research: Hang on a Seq. Advanced Genetics [Internet]. John Wiley & Sons, Ltd; 2023 [cited 2025 Feb 25];4:2200024. 10.1002/ggn2.202200024

26. Griswold AJ, Sivasankaran SK, Booven D Van, Gardner OK, Rajabli F, Whitehead PL, et al. Immune and Inflammatory Pathways Implicated by Whole Blood Transcriptomic Analysis in a Diverse Ancestry Alzheimer’s Disease Cohort. Journal of Alzheimer’s Disease [Internet]. IOS Press BV; 2020;76:1047–60. 10.3233/JAD-190855

27. Hou J, Hess JL, Zhang C, van Rooij JGJ, Hearn GC, Fan CC, et al. Meta-Analysis of Transcriptomic Studies of Blood and Six Brain Regions Identifies a Consensus of 15 Cross-Tissue Mechanisms in Alzheimer’s Disease and Suggests an Origin of Cross-Study Heterogeneity. American Journal of Medical Genetics Part B: Neuropsychiatric Genetics [Internet]. John Wiley and Sons Inc; 2025 [cited 2026 May 23];198:e33019. 10.1002/ajmg.b.33019

28. Lovestone S, Francis P, Kloszewska I, Mecocci P, Simmons A, Soininen H, et al. AddNeuroMed—The European Collaboration for the Discovery of Novel Biomarkers for Alzheimer’s Disease. Ann N Y Acad Sci [Internet]. Ann N Y Acad Sci; 2009 [cited 2025 Feb 25];1180:36–46. 10.1111/j.1749-6632.2009.05064.x

29. Frankish A, Carbonell-Sala S, Diekhans M, Jungreis I, Loveland JE, Mudge JM, et al. GENCODE: reference annotation for the human and mouse genomes in 2023. Nucleic Acids Res [Internet]. Nucleic Acids Res; 2023 [cited 2025 Feb 25];51:D942–9. 10.1093/nar/gkac1071

30. Dobin A, Davis CA, Schlesinger F, Drenkow J, Zaleski C, Jha S, et al. STAR: ultrafast universal RNA-seq aligner. Bioinformatics [Internet]. 2013;29:15–21. 10.1093/bioinformatics/bts635

31. Welsh EA, Eschrich SA, Berglund AE, Fenstermacher DA. Iterative rank-order normalization of gene expression microarray data. BMC Bioinformatics [Internet]. 2013 [cited 2025 Feb 25];14:153. 10.1186/1471-2105-14-153

32. Mcleod JC, Lim C, Stokes T, Sharif J-A, Zeynalli V, Wiens L, et al. Network-based modelling reveals cell-type enriched patterns of non-coding RNA regulation during human skeletal muscle remodelling. NAR Molecular Medicine [Internet]. Oxford Academic; 2024 [cited 2025 Feb 25];1. 10.1093/narmme/ugae016

33. Jacobs T, Jacobson SR, Fortea J, Berger JS, Vedvyas A, Marsh K, et al. The neutrophil to lymphocyte ratio associates with markers of Alzheimer’s disease pathology in cognitively unimpaired elderly people. Immun Ageing [Internet]. BioMed Central Ltd; 2024 [cited 2025 Feb 25];21:32. 10.1186/S12979-024-00435-2

34. Kikuchi M, Kobayashi K, Itoh S, Kasuga K, Miyashita A, Ikeuchi T, et al. Identification of mild cognitive impairment subtypes predicting conversion to Alzheimer’s disease using multimodal data. Comput Struct Biotechnol J [Internet]. Elsevier B.V.; 2022 [cited 2025 Feb 25];20:5296–308. 10.1016/j.csbj.2022.08.007

35. Monaco G, Lee B, Xu W, Mustafah S, Hwang YY, Carré C, et al. RNA-Seq Signatures Normalized by mRNA Abundance Allow Absolute Deconvolution of Human Immune Cell Types. Cell Rep [Internet]. 2019;26:1627–1640.e7. 10.1016/j.celrep.2019.01.041

36. Johnson WE, Li C, Rabinovic A. Adjusting batch effects in microarray expression data using empirical Bayes methods. Biostatistics [Internet]. 2007;8:118–27. 10.1093/biostatistics/kxj037

37. Tusher VG, Tibshirani R, Chu G. Significance analysis of microarrays applied to the ionizing radiation response. Proceedings of the National Academy of Sciences [Internet]. 2001;98:5116–21. 10.1073/pnas.091062498

38. Piron A, Szymczak F, Papadopoulou T, Alvelos MI, Defrance M, Lenaerts T, et al. RedRibbon: A new rank-rank hypergeometric overlap for gene and transcript expression signatures. Life Sci Alliance. Rockefeller University Press; 2023;7. 10.26508/lsa.202302203

39. Sherman BT, Hao M, Qiu J, Jiao X, Baseler MW, Lane HC, et al. DAVID: a web server for functional enrichment analysis and functional annotation of gene lists (2021 update). Nucleic Acids Res [Internet]. 2022;50:W216–21. 10.1093/nar/gkac194

40. Zhou Y, Zhou B, Pache L, Chang M, Khodabakhshi AH, Tanaseichuk O, et al. Metascape provides a biologist-oriented resource for the analysis of systems-level datasets. Nat Commun [Internet]. Nature Publishing Group; 2019 [cited 2021 Jan 21];10. 10.1038/s41467-019-09234-6

41. Timmons JA, Szkop KJ, Gallagher IJ. Multiple sources of bias confound functional enrichment analysis of global -omics data. Genome Biol [Internet]. BioMed Central Ltd.; 2015 [cited 2025 Feb 25];16:186. 10.1186/s13059-015-0761-7

42. Xu S, Hu E, Cai Y, Xie Z, Luo X, Zhan L, et al. Using clusterProfiler to characterize multiomics data. Nat Protoc [Internet]. Nature Publishing Group; 2024 [cited 2025 Feb 25];19:3292–320. 10.1038/s41596-024-01020-z

43. Fischl B. FreeSurfer. Neuroimage [Internet]. 2012;62:774–81. 10.1016/j.neuroimage.2012.01.021

44. Hanchuan Peng, Fuhui Long, Ding C. Feature selection based on mutual information criteria of max-dependency, max-relevance, and min-redundancy. IEEE Trans Pattern Anal Mach Intell [Internet]. 2005;27:1226–38. 10.1109/TPAMI.2005.159

45. Lundberg SM, Lee SI. A Unified Approach to Interpreting Model Predictions. Adv Neural Inf Process Syst [Internet]. Curran Associates, Inc.; 2017 [cited 2025 May 17]. 10.48550/arXiv.1705.07874

46. Love MI, Huber W, Anders S. Moderated estimation of fold change and dispersion for RNA-seq data with DESeq2. Genome Biology 2014 15:12 [Internet]. BioMed Central; 2014 [cited 2026 Mar 18];15:550-. 10.1186/s13059-014-0550-8

47. Zhang Y, Parmigiani G, Johnson WE. ComBat-seq: batch effect adjustment for RNA-seq count data. NAR Genom Bioinform [Internet]. Oxford Academic; 2020 [cited 2026 Mar 18];2. 10.1093/nargab/lqaa078

48. Brogan RJ, Rooyackers O, Phillips BE, Twelkmeyer B, Ross LM, Atherton PJ, et al. Biomarkers of Insulin Resistance and Their Performance as Predictors of Treatment Response in Overweight Adults. J Clin Endocrinol Metab. The Endocrine Society; 2025; 10.1210/clinem/dgaf285

49. Phillips BE, Kelly BM, Lilja M, Ponce-González JG, Brogan RJ, Morris DL, et al. A practical and time-efficient high-intensity interval training program modifies cardio-metabolic risk factors in adults with risk factors for type II diabetes. Front Endocrinol (Lausanne). Frontiers Media S.A.; 2017;8:1–11. 10.3389/fendo.2017.00229

50. Ross LM, Slentz CA, Zidek AM, Huffman KM, Shalaurova I, Otvos JD, et al. Effects of Amount, Intensity, and Mode of Exercise Training on Insulin Resistance and Type 2 Diabetes Risk in the STRRIDE Randomized Trials. Front Physiol. Frontiers Media S.A.; 2021;12. 10.3389/fphys.2021.626142

51. Sood S, Gallagher IJ, Lunnon K, Rullman E, Keohane A, Crossland H, et al. A novel multi-tissue RNA diagnostic of healthy ageing relates to cognitive health status. Genome Biol. BioMed Central Ltd.; 2015;16. 10.1186/s13059-015-0750-x

52. Li W, Shen J, Wu H, Lin L, Liu Y, Pei Z, et al. Transcriptome Analysis Reveals a Two-Gene Signature Links to Motor Progression and Alterations of Immune Cells in Parkinson’s Disease. J Parkinsons Dis [Internet]. J Parkinsons Dis; 2022 [cited 2025 Feb 25];13:25–38. 10.3233/JPD-223454

53. Muñoz-Delgado L, Macías-García D, Jesús S, Martín-Rodríguez JF, Labrador-Espinosa MÁ, Jiménez-Jaraba MV, et al. Peripheral Immune Profile and Neutrophil-to-Lymphocyte Ratio in Parkinson’s Disease. Movement Disorders [Internet]. John Wiley & Sons, Ltd; 2021 [cited 2025 Jan 30];36:2426–30. 10.1002/mds.28685

54. Nachun D, Ramos EM, Karydas A, Dokuru D, Gao F, Yang Z, et al. Systems-level analysis of peripheral blood gene expression in dementia patients reveals an innate immune response shared across multiple disorders. bioRxiv [Internet]. Cold Spring Harbor Laboratory; 2019 [cited 2026 May 22];2019.12.13.875112. 10.1101/2019.12.13.875112

55. Brodaty H, Donkin M. Family caregivers of people with dementia. Dialogues Clin Neurosci [Internet]. 2009 [cited 2026 Mar 19];11:217. 10.31887/dcns.2009.11.2/hbrodaty

56. Birkenbihl C, Salimi Y, Domingo-Fernándéz D, Lovestone S, Fröhlich H, Hofmann-Apitius M. Evaluating the Alzheimer’s disease data landscape. Alzheimer’s & Dementia: Translational Research & Clinical Interventions [Internet]. John Wiley & Sons, Ltd; 2020 [cited 2025 Jan 30];6:e12102. 10.1002/trc2.12102

57. Weiner MW, Kanoria S, Miller MJ, Aisen PS, Beckett LA, Conti C, et al. Overview of Alzheimer’s Disease Neuroimaging Initiative and future clinical trials. Alzheimer’s & Dementia [Internet]. John Wiley & Sons, Ltd; 2025 [cited 2025 Jan 30];21:e14321. 10.1002/alz.14321

58. C. Reese L, Taglialatela G. A Role for Calcineurin in Alzheimers Disease. Curr Neuropharmacol [Internet]. Bentham Science Publishers Ltd.; 2011 [cited 2026 Mar 18];9:685–92. 10.2174/157015911798376316

59. Tolias KF, Bikoff JB, Kane CG, Tolias CS, Hu L, Greenberg ME. The Rac1 guanine nucleotide exchange factor Tiam1 mediates EphB receptor-dependent dendritic spine development. Proc Natl Acad Sci U S A [Internet]. 2007 [cited 2026 Mar 18];104:7265. 10.1073/pnas.0702044104

60. Niu X, Li X, Feng Z, Han Q, Li J, Liu Y, et al. EDIL3 and VEGF Synergistically Affect Angiogenesis in Endothelial Cells. Clin Cosmet Investig Dermatol [Internet]. Dove Press; 2023 [cited 2026 Mar 18];16:1269–77. 10.2147/CCID.S411253

61. Populus, Alzheimer’s Research UK, MSD. Detecting and diagnosing Alzheimer’s disease: Enhancing our understanding of public attitudes to improving early detection and diagnosis [Internet]. Cambridge, UK; 2019. https://www.alzheimersresearchuk.org/wp-content/uploads/2019/12/1132267-Public-Perceptions-Report_v5.pdf. Accessed 30 Jan 2025

62. Xu KY, Violich I, Hutchins E, Alsop E, Nalls MA, Moore A, et al. Decreased SNCA expression in whole-blood RNA analysis of Parkinson’s disease adjusting for neutrophils. npj Parkinson’s Disease 2025 11:1 [Internet]. Nature Publishing Group; 2025 [cited 2026 Mar 18];11:292-. 10.1038/s41531-025-01062-4

63. Riley LK, Rupert J. Evaluation of Patients with Leukocytosis. Am Fam Physician [Internet]. 2015;92:1004–11. https://www.aafp.org/pubs/afp/issues/2015/1201/p1004.html

64. Forget P, Khalifa C, Defour J-P, Latinne D, Van Pel M-C, De Kock M. What is the normal value of the neutrophil-to-lymphocyte ratio? BMC Res Notes [Internet]. BioMed Central; 2017 [cited 2025 Jan 30];10:12. 10.1186/s13104-016-2335-5

65. Petrone AB, Eisenman RD, Steele KN, Mosmiller LT, Urhie O, Zdilla MJ. Temporal dynamics of peripheral neutrophil and lymphocytes following acute ischemic stroke. Neurological Sciences [Internet]. Springer-Verlag Italia s.r.l.; 2019 [cited 2025 Jan 30];40:1877–85. 10.1007/s10072-019-03919-y

66. Koyama A, O’Brien J, Weuve J, Blacker D, Metti AL, Yaffe K. The Role of Peripheral Inflammatory Markers in Dementia and Alzheimer’s Disease: A Meta-Analysis. J Gerontol A Biol Sci Med Sci [Internet]. Oxford Academic; 2013 [cited 2025 Sep 26];68:433–40. 10.1093/gerona/gls187

67. Soehnlein O, Steffens S, Hidalgo A, Weber C. Neutrophils as protagonists and targets in chronic inflammation. Nat Rev Immunol [Internet]. Nature Publishing Group; 2017 [cited 2025 Jan 30];17:248–61. 10.1038/nri.2017.10

68. Pietronigro EC, Della Bianca V, Zenaro E, Constantin G. NETosis in Alzheimer’s Disease. Front Immunol [Internet]. Frontiers Research Foundation; 2017 [cited 2025 Sep 30];8:228493. 10.3389/fimmu.2017.00211

69. Aries ML, Hensley-McBain T. Neutrophils as a potential therapeutic target in Alzheimer’s disease. Front Immunol [Internet]. Frontiers Media S.A.; 2023 [cited 2025 Jan 30];14:1123149. 10.3389/fimmu.2023.1123149

70. Dong Y, Lagarde J, Xicota L, Corne H, Chantran Y, Chaigneau T, et al. Neutrophil hyperactivation correlates with Alzheimer’s disease progression. Ann Neurol [Internet]. John Wiley & Sons, Ltd; 2018 [cited 2025 Sep 30];83:387–405. 10.1002/ana.25159

71. Seda V, Mraz M. B-cell receptor signalling and its crosstalk with other pathways in normal and malignant cells. Eur J Haematol [Internet]. John Wiley & Sons, Ltd; 2015 [cited 2025 Feb 14];94:193–205. 10.1111/ejh.12427

72. Gómez Hernández G, Morell M, Alarcón-Riquelme ME. The Role of BANK1 in B Cell Signaling and Disease. Cells [Internet]. Multidisciplinary Digital Publishing Institute; 2021 [cited 2025 Feb 14];10:1184. 10.3390/cells10051184

73. Jorfi M, Maaser-Hecker A, Tanzi RE. The neuroimmune axis of Alzheimer’s disease. Genome Med [Internet]. BioMed Central Ltd; 2023 [cited 2025 Apr 16];15:6. 10.1186/s13073-023-01155-w

74. Feng W, Zhang Y, Ding S, Chen S, Wang T, Wang Z, et al. B lymphocytes ameliorate Alzheimer’s disease-like neuropathology via interleukin-35. Brain Behav Immun [Internet]. Academic Press; 2023 [cited 2025 Feb 12];108:16–31. 10.1016/j.bbi.2022.11.012

75. Kim K, Wang X, Ragonnaud E, Bodogai M, Illouz T, DeLuca M, et al. Therapeutic B-cell depletion reverses progression of Alzheimer’s disease. Nat Commun [Internet]. Nature Research; 2021 [cited 2025 Feb 12];12:2185. 10.1038/s41467-021-22479-4

76. Alzheimer’s Association. 2024 Alzheimer’s disease facts and figures [Internet]. Alzheimer’s & Dementia. John Wiley & Sons, Ltd; 2024 May. 10.1002/alz.13809

77. Lanctôt KL, Hahn-Pedersen JH, Eichinger CS, Freeman C, Clark A, Tarazona LRS, et al. Burden of Illness in People with Alzheimer’s Disease: A Systematic Review of Epidemiology, Comorbidities and Mortality. Journal of Prevention of Alzheimer’s Disease. Serdi-Editions; 2024;11:97–107. 10.14283/jpad.2023.61

78. Jonsson A, Isomaa B, Tuomi T, Taneera J, Salehi A, Nilsson P, et al. A Variant in the KCNQ1 Gene Predicts Future Type 2 Diabetes and Mediates Impaired Insulin Secretion. Diabetes [Internet]. American Diabetes Association; 2009 [cited 2026 Mar 18];58:2409–13. 10.2337/db09-0246

79. Zhou X, Zhu J, Bao Z, Shang Z, Wang T, Song J, et al. A variation in KCNQ1 gene is associated with repaglinide efficacy on insulin resistance in Chinese Type 2 Diabetes Mellitus Patients. Scientific Reports 2016 6:1 [Internet]. Nature Publishing Group; 2016 [cited 2026 Mar 18];6:37293-. 10.1038/srep37293

80. Sithu SD, Malovichko M V., Riggs KA, Wickramasinghe NS, Winner MG, Agarwal A, et al. Atherogenesis and metabolic dysregulation in LDL receptor–knockout rats. JCI Insight [Internet]. American Society for Clinical Investigation; 2017 [cited 2026 Mar 18];2:e86442. 10.1172/jci.insight.86442

81. Mulvihill EE, Allister EM, Sutherland BG, Telford DE, Sawyez CG, Edwards JY, et al. Naringenin Prevents Dyslipidemia, Apolipoprotein B Overproduction, and Hyperinsulinemia in LDL Receptor–Null Mice With Diet-Induced Insulin Resistance. Diabetes [Internet]. American Diabetes Association; 2009 [cited 2026 Mar 18];58:2198–210. 10.2337/db09-0634

82. Bonfleur ML, Vanzela EC, Ribeiro RA, de Gabriel Dorighello G, de França Carvalho CP, Collares-Buzato CB, et al. Primary hypercholesterolaemia impairs glucose homeostasis and insulin secretion in low-density lipoprotein receptor knockout mice independently of high-fat diet and obesity. Biochimica et Biophysica Acta (BBA) - Molecular and Cell Biology of Lipids [Internet]. Elsevier; 2010 [cited 2026 Mar 18];1801:183–90. 10.1016/j.bbalip.2009.10.012

83. Fujimoto BA, Young M, Carter L, Pang APS, Corley MJ, Fogelgren B, et al. The exocyst complex regulates insulin-stimulated glucose uptake of skeletal muscle cells. American Journal of Physiology-Endocrinology and Metabolism [Internet]. American Physiological Society Bethesda, MD ; 2019 [cited 2026 Apr 27];317:E957–72. 10.1152/ajpendo.00109.2019

84. Chandra NC. A comprehensive account of insulin and LDL receptor activity over the years: A highlight on their signaling and functional role. J Biochem Mol Toxicol [Internet]. John Wiley and Sons Inc; 2021 [cited 2026 Mar 18];35:e22840. 10.1002/jbt.22840

85. Qaisar N, Lin S, Ryan G, Yang C, Oikemus SR, Brodsky MH, et al. A Critical Role for the Type I Interferon Receptor in Virus-Induced Autoimmune Diabetes in Rats. Diabetes [Internet]. American Diabetes Association; 2017 [cited 2026 Mar 18];66:145–57. 10.2337/db16-0462

86. Alwazzan O, Khan A, Patras I, Slabaugh G. MOAB: Multi-Modal Outer Arithmetic Block for Fusion of Histopathological Images and Genetic Data for Brain Tumor Grading. 2023 IEEE 20th International Symposium on Biomedical Imaging (ISBI) [Internet]. IEEE; 2023. p. 1–5. 10.1109/ISBI53787.2023.10230698

87. Stahlschmidt SR, Ulfenborg B, Synnergren J. Multimodal deep learning for biomedical data fusion: a review. Brief Bioinform [Internet]. Oxford Academic; 2022;23. 10.1093/bib/bbab569

88. Maddalena L, Granata I, Giordano M, Manzo M, Guarracino MR. Integrating Different Data Modalities for the Classification of Alzheimer’s Disease Stages. SN Comput Sci [Internet]. 2023;4:249. 10.1007/s42979-023-01688-2

