## Supplementary Figures for "Sex and insulin resistance biomarker modelling in a new large-scale Alzheimer’s disease transcriptomic resource"

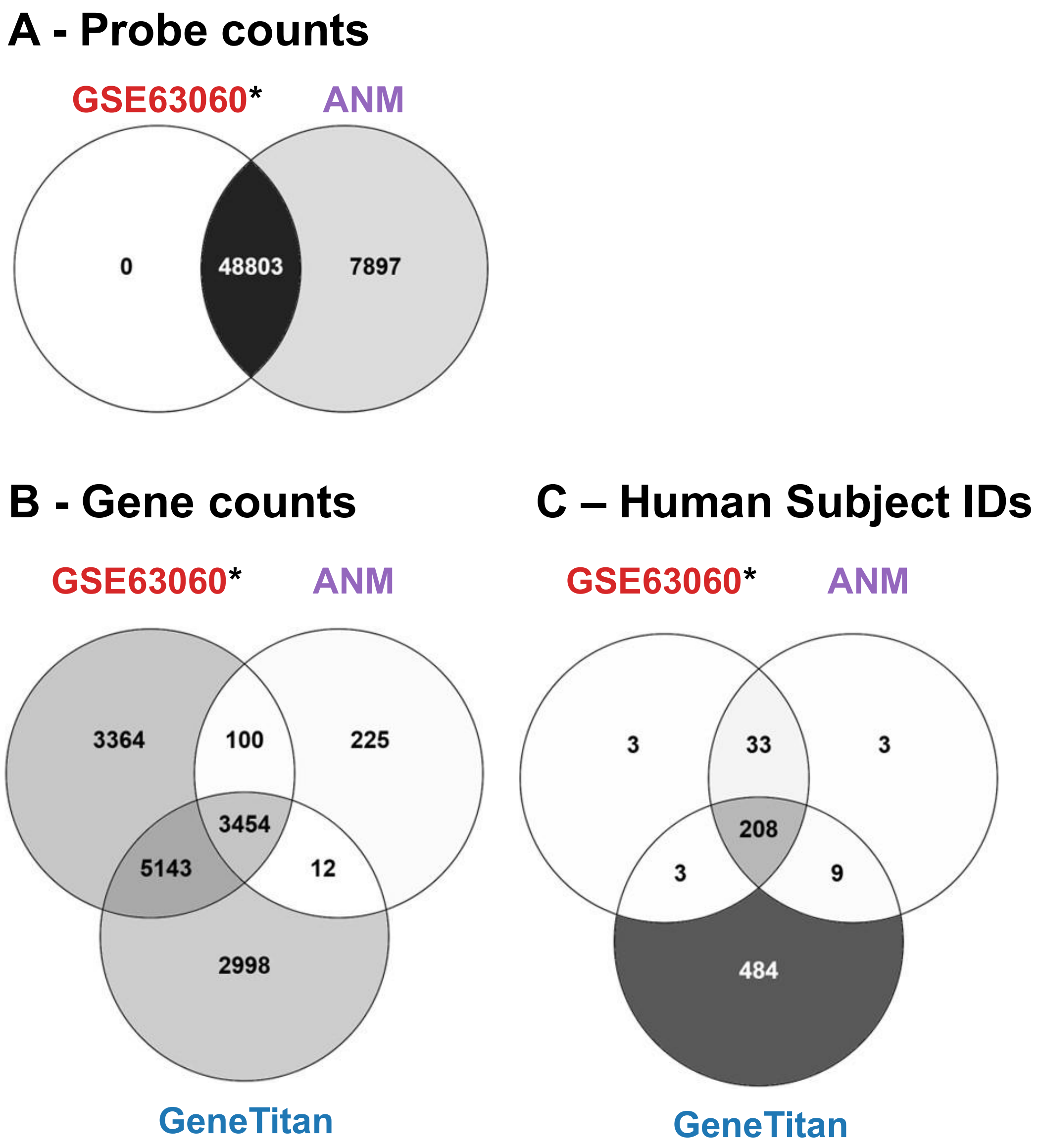


**Figure S1. GeneTitan Metrics for probes or genes (rows of data) per data set. *GSE63060 raw data is largely identical as it comes from the same laboratory files used by ANMerge, but was realigned and annotated to the latest genome in 2024.** The third data set was produced on an Affymetrix GeneTitan (GeneTitan) using an HTHGU133Plus PM array. **A:** Venn diagrams comparing Illumina probes in GSE63060 after full processing and realignment to the current genome/transcriptome versus the Illumina batch 1 data deposited by ANMerge (ANM). **B:** Ensembl gene IDs (ENSG) present after processing and realignment to the current genome/transcriptome processed in the GSE6030 data, the ANM data and new Affymetrix GeneTitan data (also processed and re-aligned to the current genome) and **C:** Total number of AD and CTL subjects (Final Diagnosis labels) available in each data set and the overlap in membership across the three sources of data (GSE63060: n=247, ANM: n=253 and Affymetrix: n=704 individuals (1021 arrays in total as we produced technical duplicates)). Note that ANMerge has additional transcriptomic data from AddNeuroMed batch 2 (unnormalized AD + CTL, n=253), but with far fewer accompanying MRI scans (n=7).

A

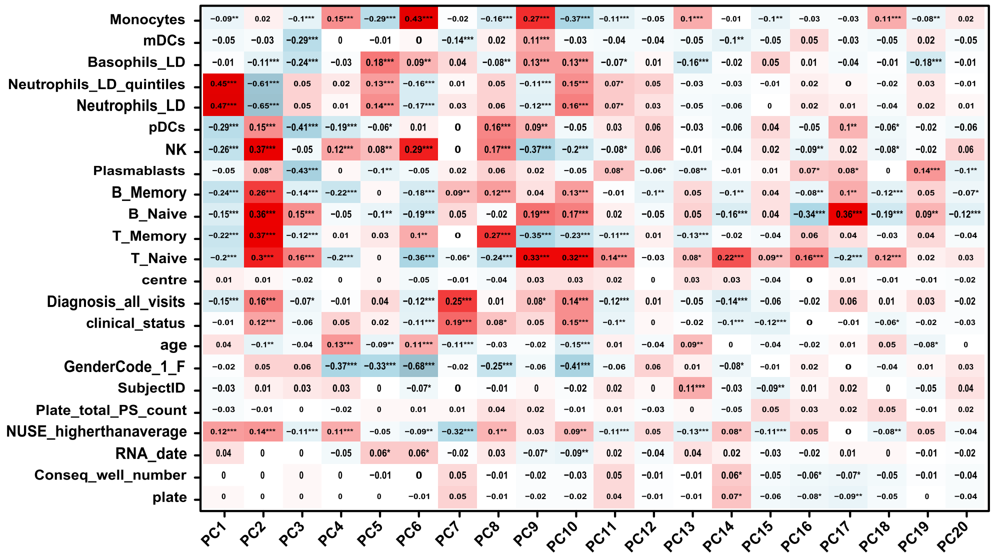


B

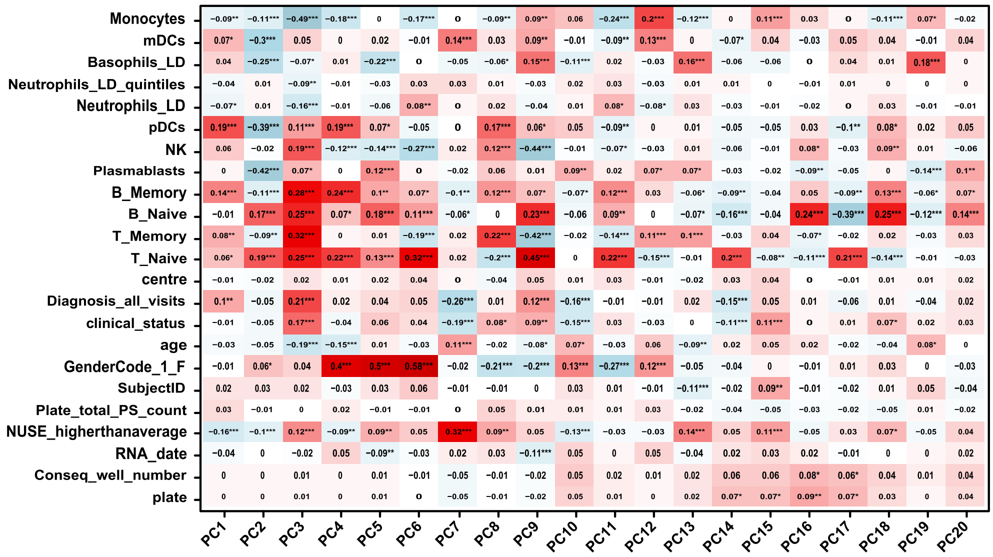


**Figure S2. Principal component (PC) analysis of lab and biological variables in Affymetrix GeneTitan transcriptomics dataset (N = 1021).** Asterisks indicate levels of statistical significance: *P* value < 0.05 (*), *P* value < 0.01 (**), *P* value < 0.001 (***). Colour gradient ranges from blue, indicating strong negative values, to red, indicating strong positive values. **A:** PC analysis in the whole blood data (corrected for plate total signal and clinical centre adjusted data). Note that Age, Diagnosis and Sex are still clearly visible as related to PCs. **B:** PC analysis after combat adjustment for variation in neutrophil quintiles limits the influence of neutrophil count on variation in gene expression. Note that estimated neutrophil counts covary with other cell types due to the deconvolution method (which we scale to a total cell count of 100). Abbreviations: PC, principal component

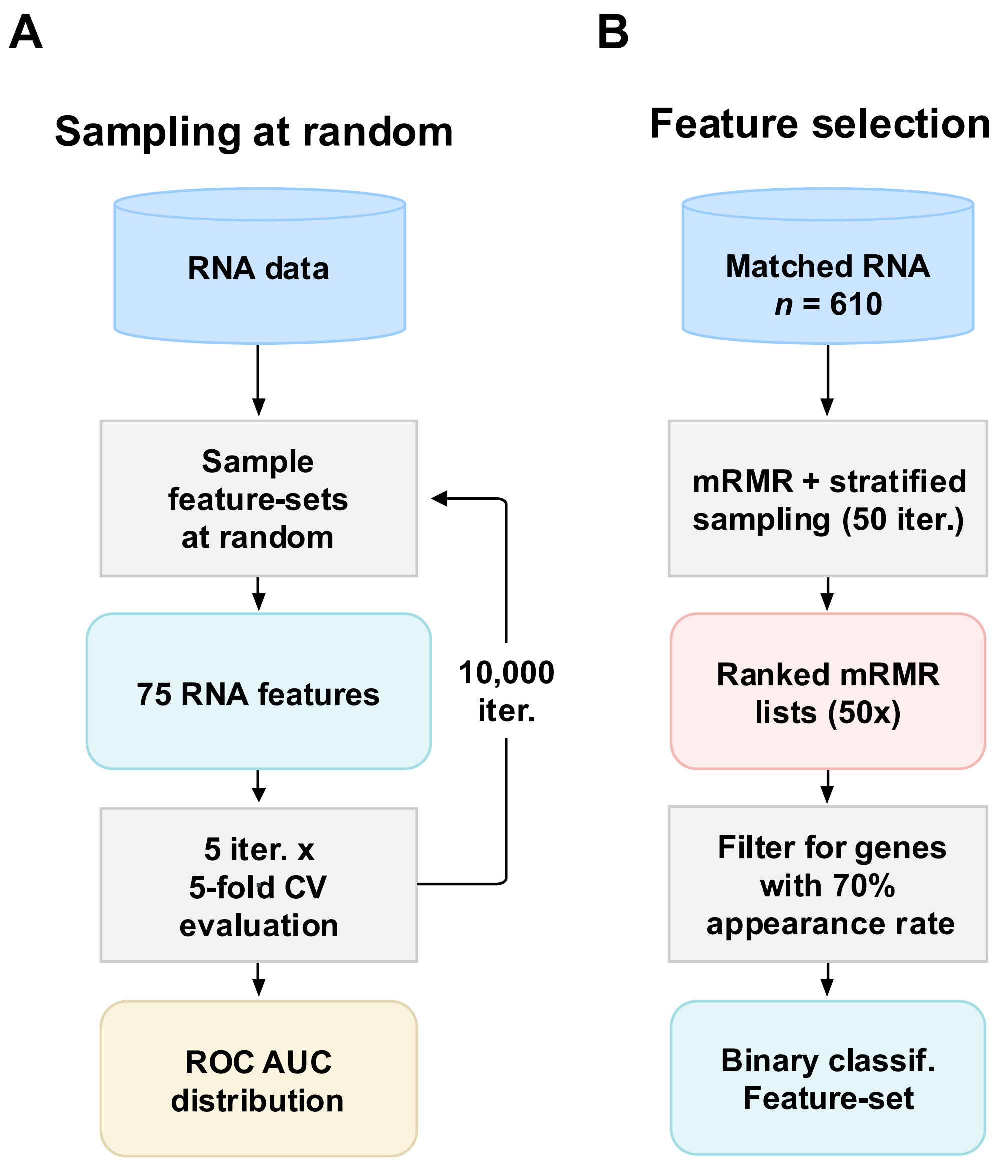


**Figure S3. The workflow for evaluating systematic bias in AD and CTL blood transcriptome profiles and feature selection strategy. A:** The workflow shows that 75 transcriptomics features were sampled at random from all available transcriptomics features in each data source. These features were used to train logistic regression classifiers for AD vs. CTL classification, using 5 repeats of 5-fold cross-validation. This process was repeated for 10,000 iterations to obtain a distribution of AUC scores. **B:** mRMR feature selection was applied 50 times on randomly selected 80% stratified subsamples of subjects from the matched transcriptomics data (n=610) to create 50 rank order mRMR lists of the top 100 transcriptomics features ('genes'). The top 100 genes from each of the 50 mRMR analyses were ranked, and the average score for each gene was used to create a single overall rank. This rank-ordered list is then filtered for genes which appear in >=70% of the 50 mRMR runs to determine the final classification feature-set. Abbreviations: mRMR, minimum Redundancy Maximum Relevance; AD, Alzheimer's disease; CTL, Control; AUC, area under the receiver operating characteristic curve.

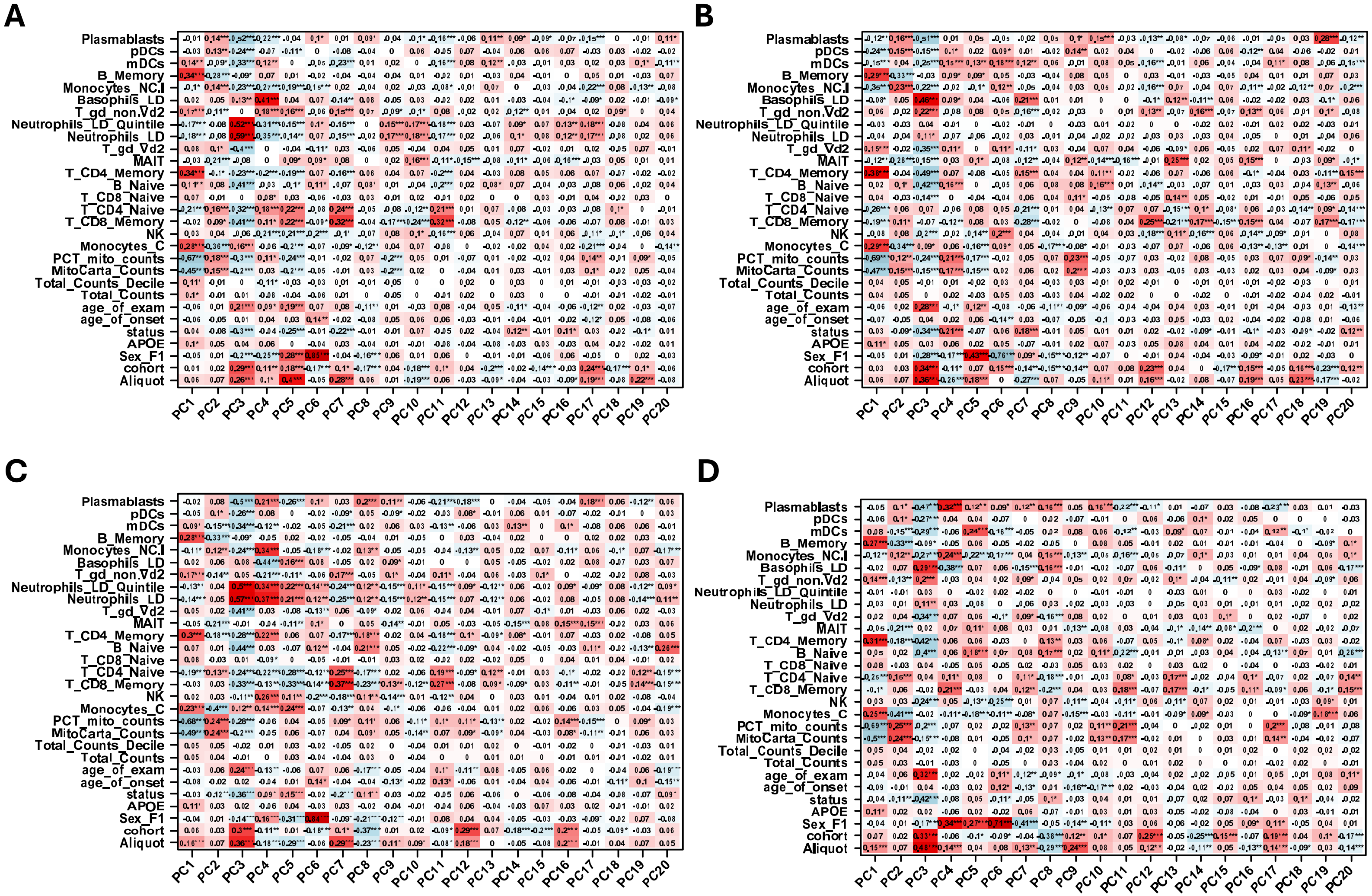


**Figure S4. Principal component (PC) analysis of lab and biological variables in AD RNAseq dataset (N=562).** Asterisks indicate levels of statistical significance: *P* value < 0.05 (*), *P* value < 0.01 (**), *P* value < 0.001 (***). Colour gradient ranges from blue, indicating strong negative values, to red, indicating strong positive values.

**Figure S4 (Continued). A**: PC analysis in standard aligned whole blood RNAseq data (corrected for total counts), **B**: PC analysis in standard aligned whole blood RNAseq data after ComBat-SEQ adjustment for variation in neutrophil quintiles limits the influence of neutrophil count on variation in gene expression, **C**: PC analysis in GeneTitan GTF-aligned whole blood RNAseq data (corrected for total counts), **D**: PC analysis in GeneTitan GTF-aligned whole blood RNAseq data after ComBat-SEQ adjustment for variation in neutrophil quintiles. Abbreviations: AD, Alzheimer's disease; PC, principal component; RNA-seq, RNA sequencing; GTF, gene transfer format.

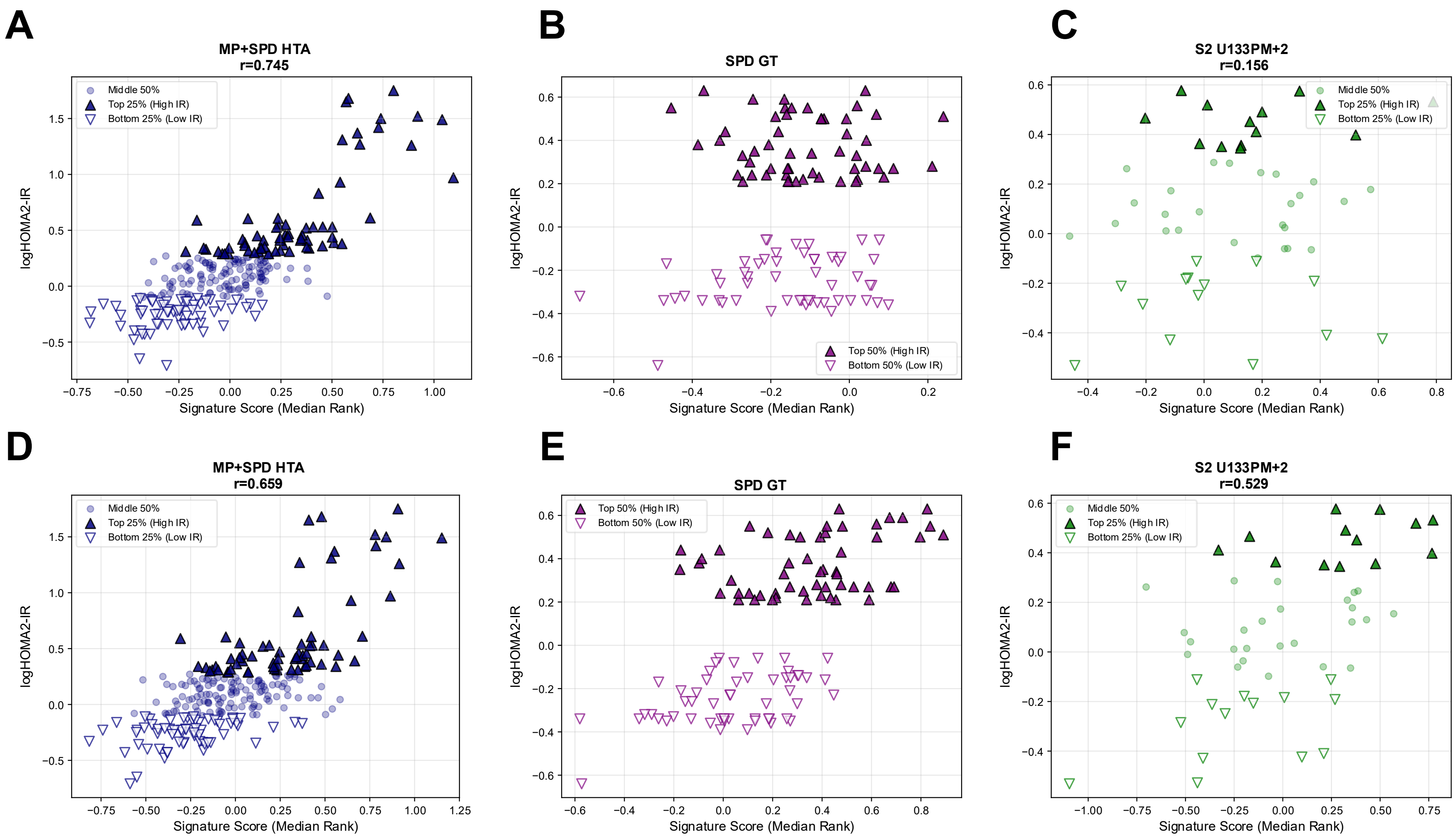


**Figure S5. Validation of insulin resistance (IR) rank gene scores in independent metabolic array cohorts.** Spearman correlation between the initial 147-gene IR rank gene score and log2 HOMA2-IR in **A:** the combined MP+SPD development cohort, **B:** the SPD GeneTitan validation cohort, **C:** the S2 U133PM validation cohort, with points colored by IR group.

**Figure S5 (continued).** In the development cohort, rank gene scores were strongly correlated with log2 HOMA2-IR (**A**, Spearman rho = 0.745, P < 0.001). In the SPD GeneTitan data, the signature visibly segregated the two groups into high and low HOMA2-IR status (**B**), while for S2 U133PM (**C**, rho = 0.156, *P* = 0.269), the rank performance was weak. **E-F:** Performance of the filtered 62-gene cross-platform IR signature, restricted to genes with concordant direction of association with log2 HOMA2-IR across all three cohorts. Using this filtered signature, rank gene scores remained strongly correlated with log2 HOMA2-IR in the combined MP+SPD cohort (**D**, rho = 0.659, P < 0.001). In SPD GeneTitan (**E**), the signature robustly separated the two IR populations, and in S2 U133PM (**F**, rho = 0.438, P < 0.001), it demonstrated improved generalisability. Abbreviations: IR, Insulin resistance; HOMA2-IR, Homeostatic Model Assessment 2 of Insulin Resistance; MP, META-PREDICT; SPD, STRRIDE-PD; S2, STRRIDE-AT/RT; HTA, Human Transcriptome Array; GT, GeneTitan; U133PM, Affymetrix Human Genome U133 Plus 2.0 microarray.

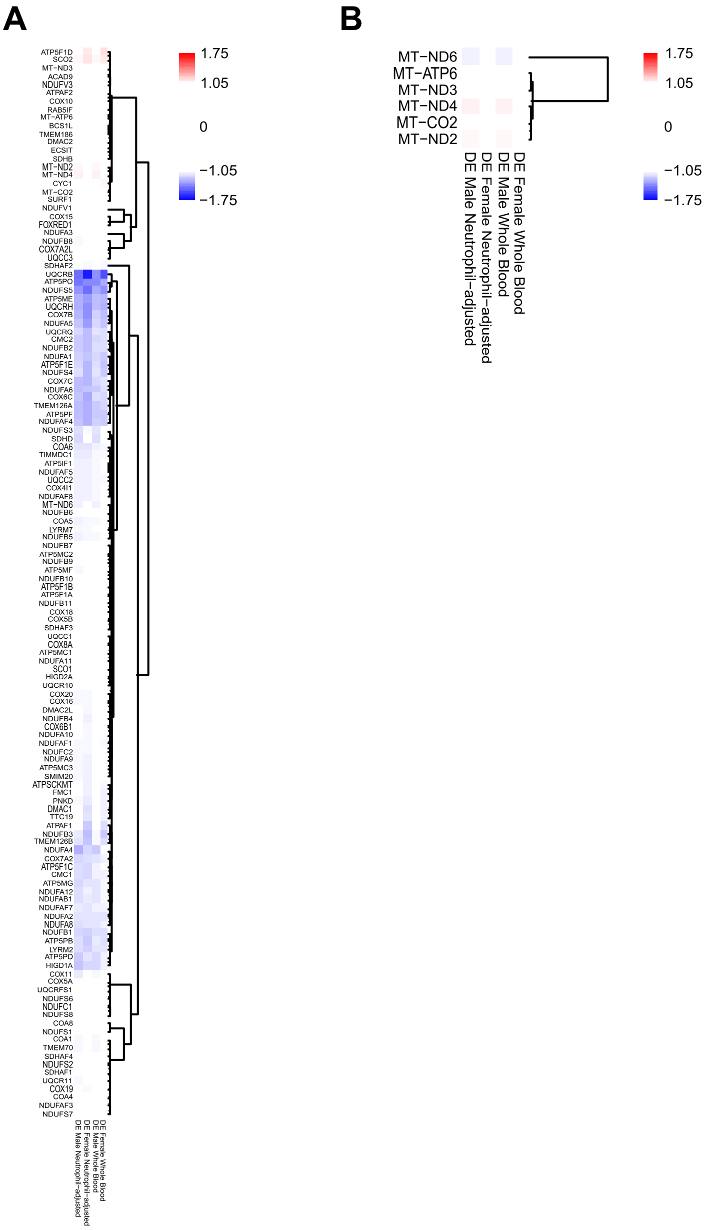


**Figure S6**. **Using the gene-sets identified by the Broad Institute MitoCarta database, we replotted the DE values as heatmaps (core oxidative phosphorylation (OXPHOS) genes and genes encoded by the mitochondrial DNA (mtDNA).** Each plot shows DE in male whole blood, female whole blood, male neutrophil-adjusted blood, and female neutrophil-adjusted blood. **A:** Nuclear-encoded OXPHOS genes. **B:** mtDNA genes. We find no evidence of altered mtDNA-encoded transcript expression in AD blood. Abbreviations: DE, Differential expression; OXPHOS, oxidative phosphorylation; mtDNA, mitochondrial DNA.

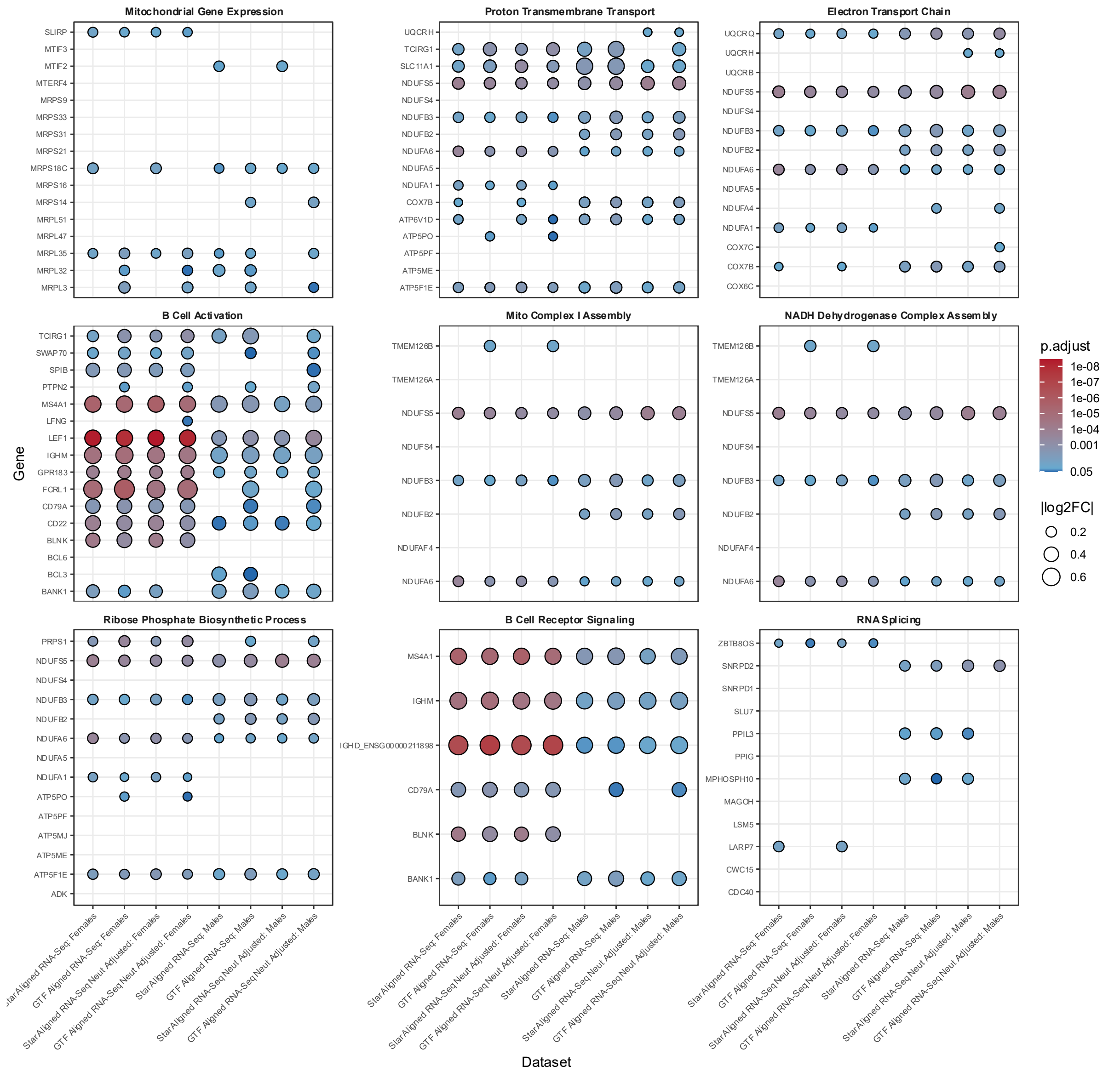


**Figure S7. Differential expression dot plots across Female and Male AD blood RNAseq datasets for genes from biological processes identified in the AD GeneTitan data (see Figure 3).** Each panel shows one GO term, with rows representing individual genes and columns representing AD case-control data from female (n=361, n(AD)=233, n(CTL)=118) and male (n=197, n(AD)=129, n(CTL)=68) subjects, analysed with DESeq2. Dot size indicates the absolute log2 fold change (|log2FC|) for each gene in that dataset, and dot colour indicates the Benjamini–Hochberg–adjusted P value (red = more significant, blue = less significant). Abbreviations: AD, Alzheimer's disease; CTL, healthy control; GO, Gene ontology

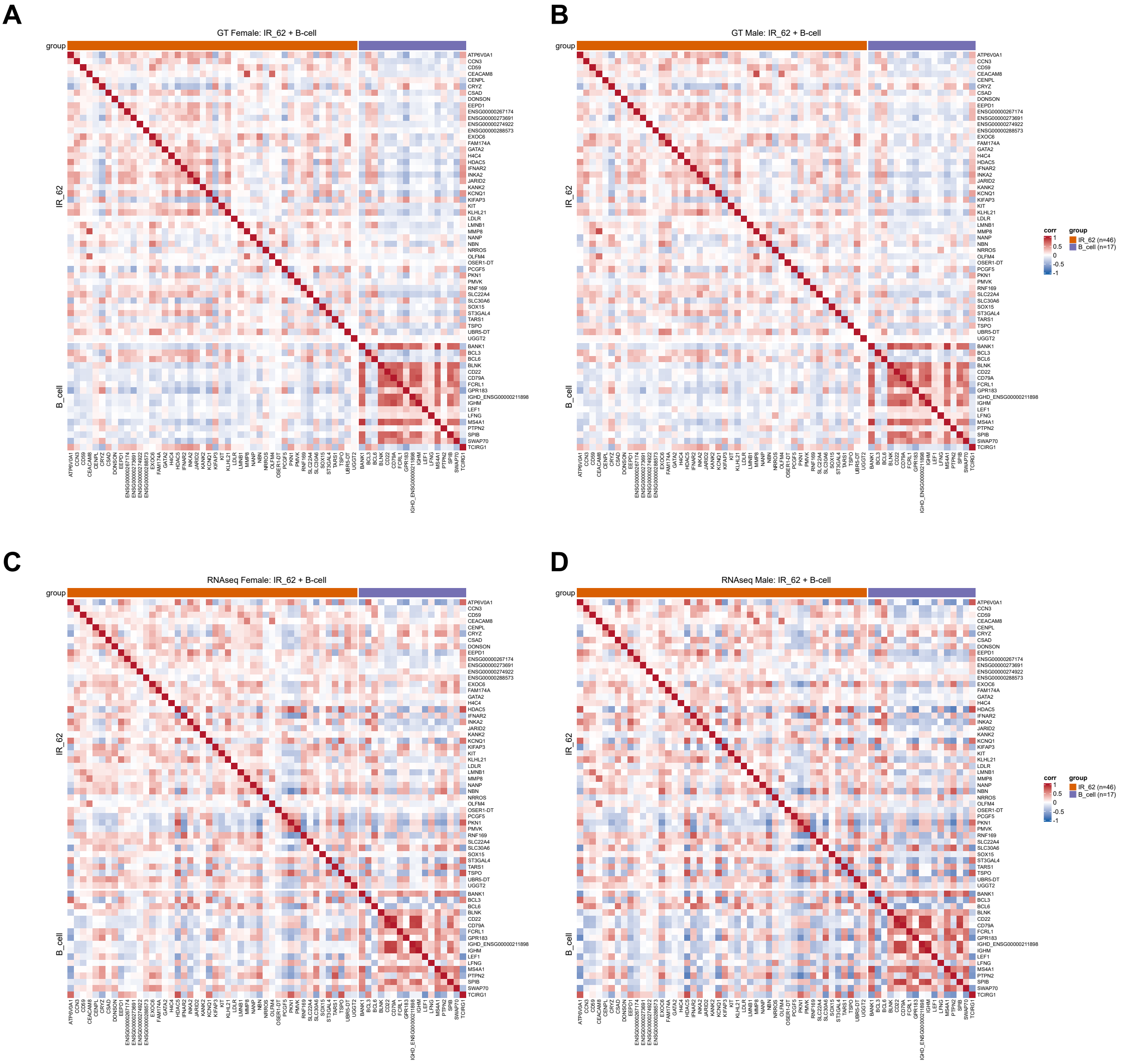


**Figure S8. Correlation heatmap of IR signature and enriched B-cell genes in whole-blood data.** Pearson correlation coefficients between genes in both the cross-platform IR signature and B-cell genes identified from DE clusterprofiler analyses on GeneTitan array data were computed in women and men in the GeneTitan and RNA-seq datasets. Only genes detected in both datasets were included (IR signature, n=46; B-cell genes, n=17). **A:** GeneTitan Female. **B:** GeneTitan Male. **C:** RNAseq Female. **D:** RNAseq Male. Abbreviations: IR, Insulin resistance; GT, GeneTitan array

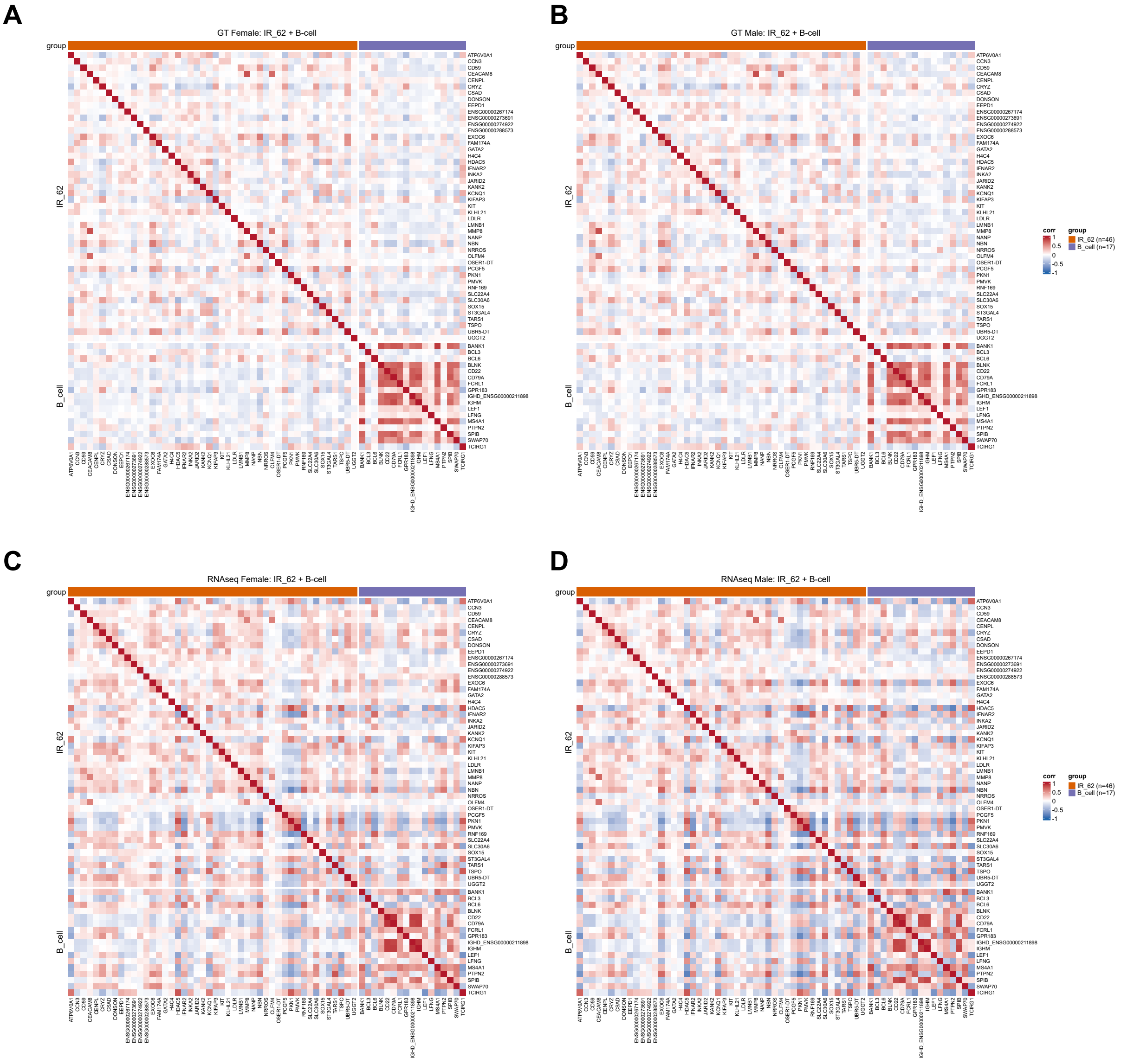


**Figure S9. Correlation heatmap of IR signature and enriched B-cell genes in neutrophil-adjusted data.** Pearson correlation coefficients between genes in both the cross-platform IR signature and B-cell genes identified from DE clusterprofiler analyses on GeneTitan array data were computed in women and men in the GeneTitan and RNA-seq datasets. Only genes detected in both datasets were included (IR signature, n=46; B-cell genes, n=17). **A:** GeneTitan Female. **B:** GeneTitan Male. **C:** RNAseq Female. **D:** RNAseq Male. Abbreviations: IR, Insulin resistance; GT, GeneTitan array

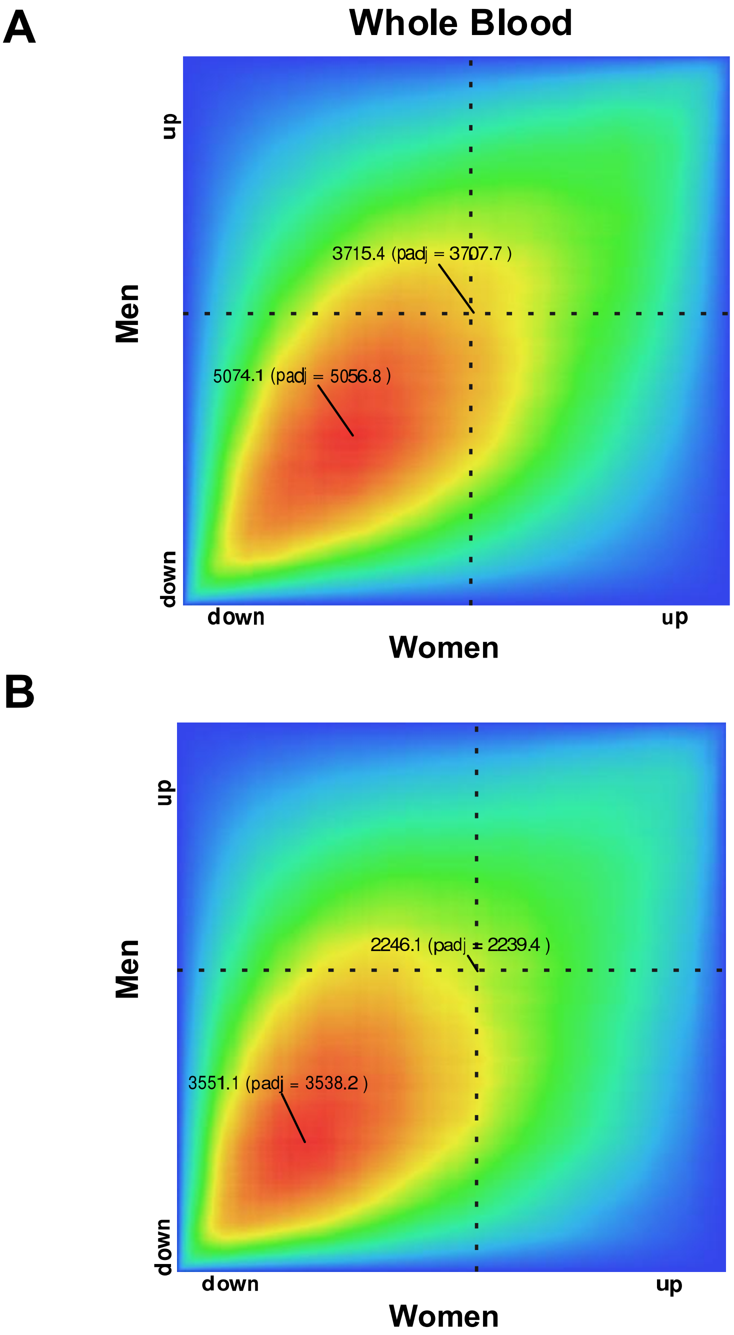


**Figure S10. Examination of the consistency of differential gene expression (AD vs CTL) in men versus women, using the RedRibbon R Package.** This ranking order method, relying on the hypergeometric distribution, allows us to directly compare the difference in gene expression in women (Control vs AD) with the difference in gene expression in men (Control vs AD), without applying a hard threshold. This is a more sensitive approach for evaluating whether two groups show consistency in biological response (compared with hard thresholding and Venn diagram-based membership analysis). **A:** GeneTitan whole blood gene expression corrected for technical variance, and **B:** GeneTitan Neutrophil and technical-adjusted gene expression (See methods).

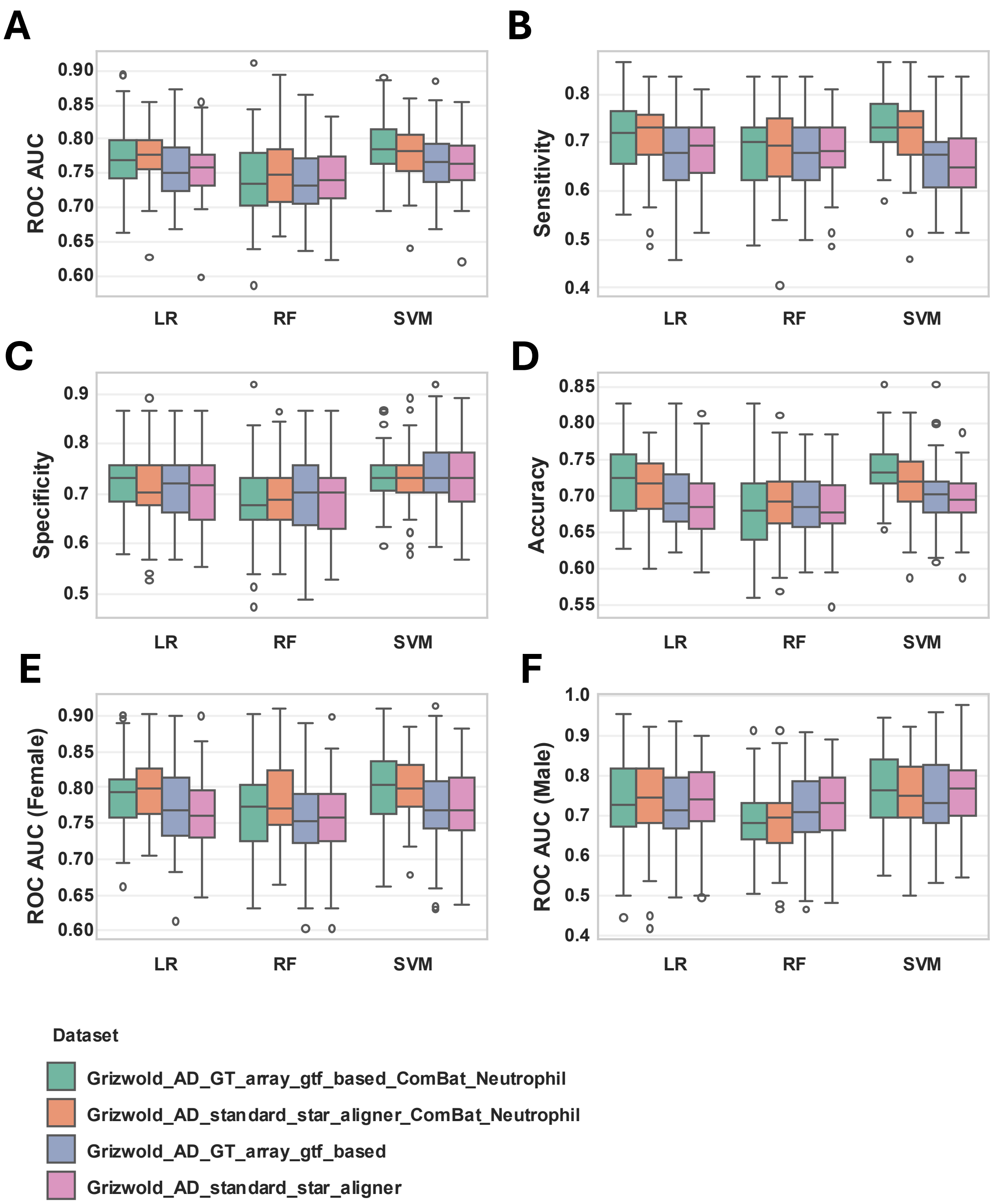


**Figure S11. Performance of binary Alzheimer's disease (AD) versus control (CTL) classifiers derived from AD whole-blood array signatures when applied to an independent AD RNA-seq cohort, with and without adjustment for deconvoluted neutrophil fraction (n = 372; 186 AD, 186 CTL).** Panels **A–F** show boxplots of cross-validated receiver operating characteristic area under the curve (ROC AUC; A, overall; E, females; F, males), sensitivity (B), specificity (C) and overall accuracy (D) for three machine-learning algorithms (logistic regression, LR; random forest, RF; support vector machine, SVM). Classifier performance was estimated using 10 iterations of 5-fold stratified cross-validation, and boxplots summarise the distribution of metrics across all resampling iterations.

**Figure S11 (Continued).** For each algorithm and metric, performance is shown for models trained on the original whole-blood AD array signature or on the neutrophil-adjusted signature and evaluated in RNA-seq data processed with four alternative pipelines (Grizwold AD GT array gtf-based, Grizwold AD standard star aligner, and data with further neutrophil adjustment using ComBat-Seq). Abbreviations: AD, Alzheimer's disease; CTL, control; LR, logistic regression; RF, random forest; AUC, area under the receiver operating characteristic curve; SVM, support vector machine.

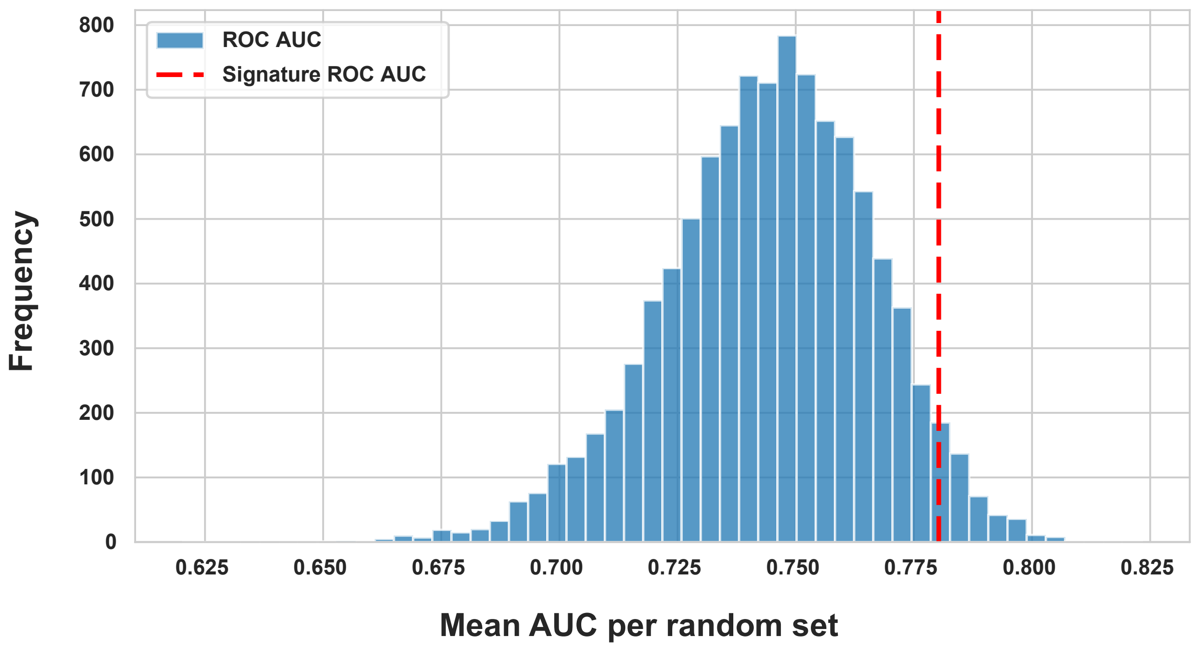


**Figure S12. Performance of SVC models trained on neutrophil-adjusted AD array blood signature versus random gene sets in AD GeneTitan GTF-aligned RNA-seq.** Distribution of mean AUC values for support vector classifiers trained on 10,000 random 39-gene sets, evaluated on GeneTitan GTF-aligned RNA-seq data from AD and control subjects using neutrophil-adjusted expression values and 10 iterations of stratified 5-fold cross-validation (50 test folds per iteration). The vertical line marks the mean AUC of the 39-gene neutrophil-adjusted AD array signature under the same cross-validation procedure. The empirical P value was estimated as the proportion of random gene-set AUCs greater than or equal to the signature AUC. AD, Alzheimer’s disease; AUC, area under the receiver operating characteristic curve; GTF, Gene Transfer Format; SVC, support vector classifier.

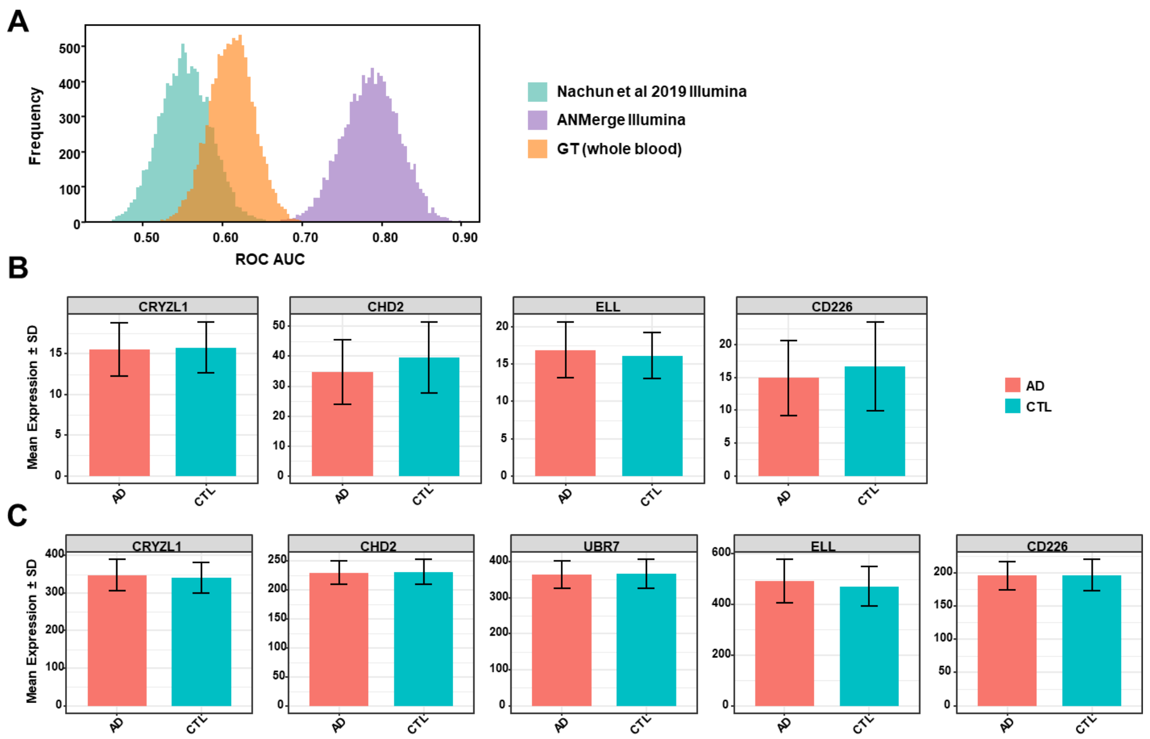


**Figure S13. Evaluation of an Illumina whole-blood AD cohort and reported marker genes. A:** Distribution of receiver operating characteristic (ROC) area under the curve (AUC) values from the sampling-at-random procedure applied to the batch-corrected Illumina GSE140829 provided by Nachun et al (AD and controls only, n = 427), our new GeneTitan (GT) whole-blood data (n = 610) and the ANMerge re-processed Illumina data. **B:** Mean expression ± SD of the top AD DE genes reported in the recent AD transcriptome meta-analysis study (CRYZL1, CHD2, ELL, CD226, see Results) in the GT whole-blood cohort (n = 1,021; AD vs CTL). **C:** Mean expression ± SD of the same genes, and UBR7 (meta-analysis DEG not detected in GT data) in the processed Illumina GSE140829 matrix (n = 427; AD vs CTL), illustrating near-random classification performance and minimal AD–CTL differences for these markers. Abbreviations: AD, Alzheimer's disease; ROC, receiver operating characteristic; AUC, area under the curve; SD, standard deviation; DE, differentially expressed; CTL, control; GT, GeneTitan.

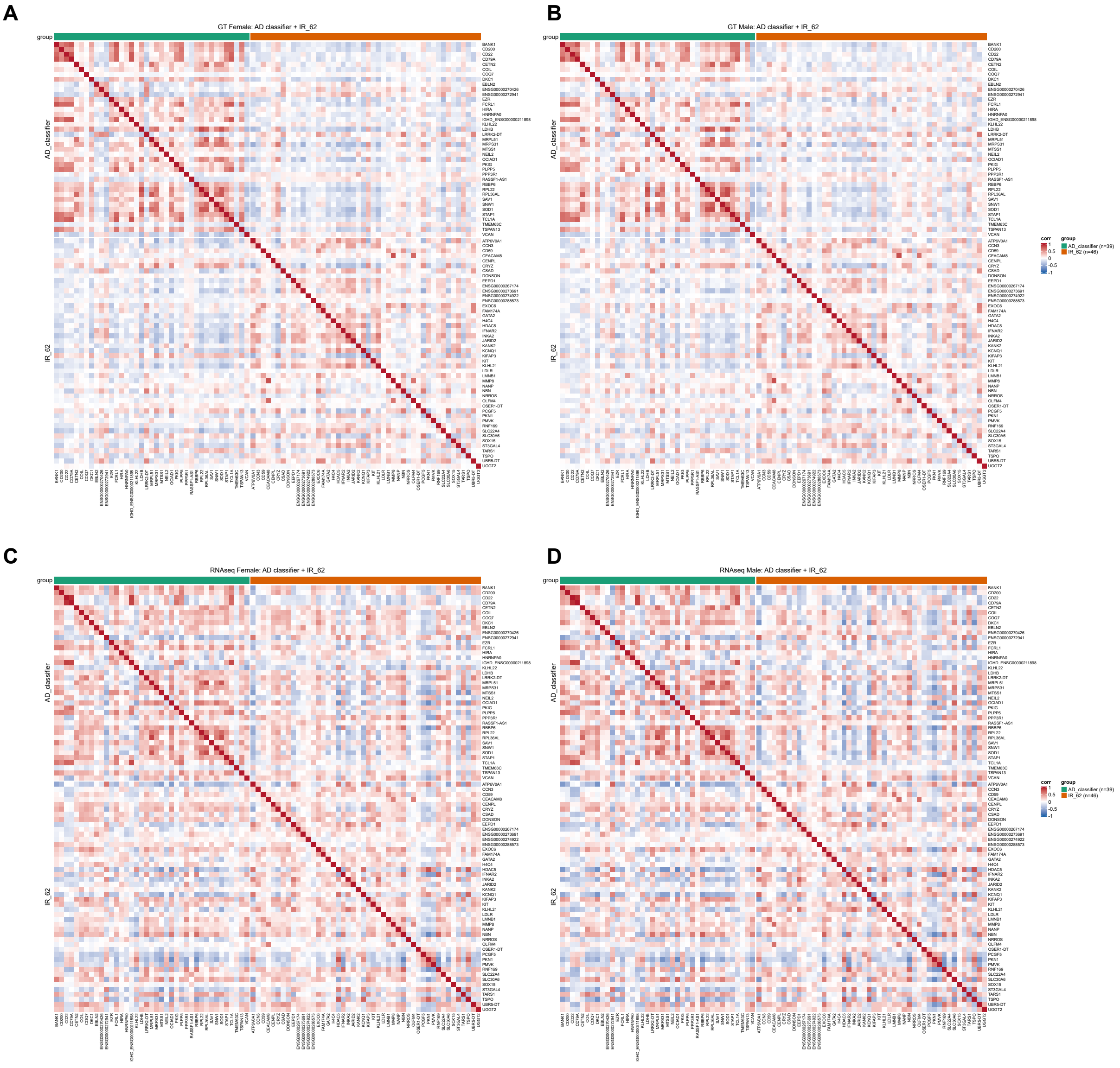


**Figure S14. Correlation heatmap of IR signature and AD classification signature genes in whole-blood data.** Pearson correlation coefficients between genes in both the whole-blood AD classification array signature and cross-platform IR signature were computed in women and men in the GeneTitan and RNA-seq datasets. Only genes detected in both datasets were included (AD classification signature, n=39; IR signature, n=46). **A:** GeneTitan Female. **B:** GeneTitan Male. **C:** RNAseq Female. **D:** RNAseq Male. Abbreviations: AD, Alzheimer's disease; IR, Insulin resistance; GT, GeneTitan array

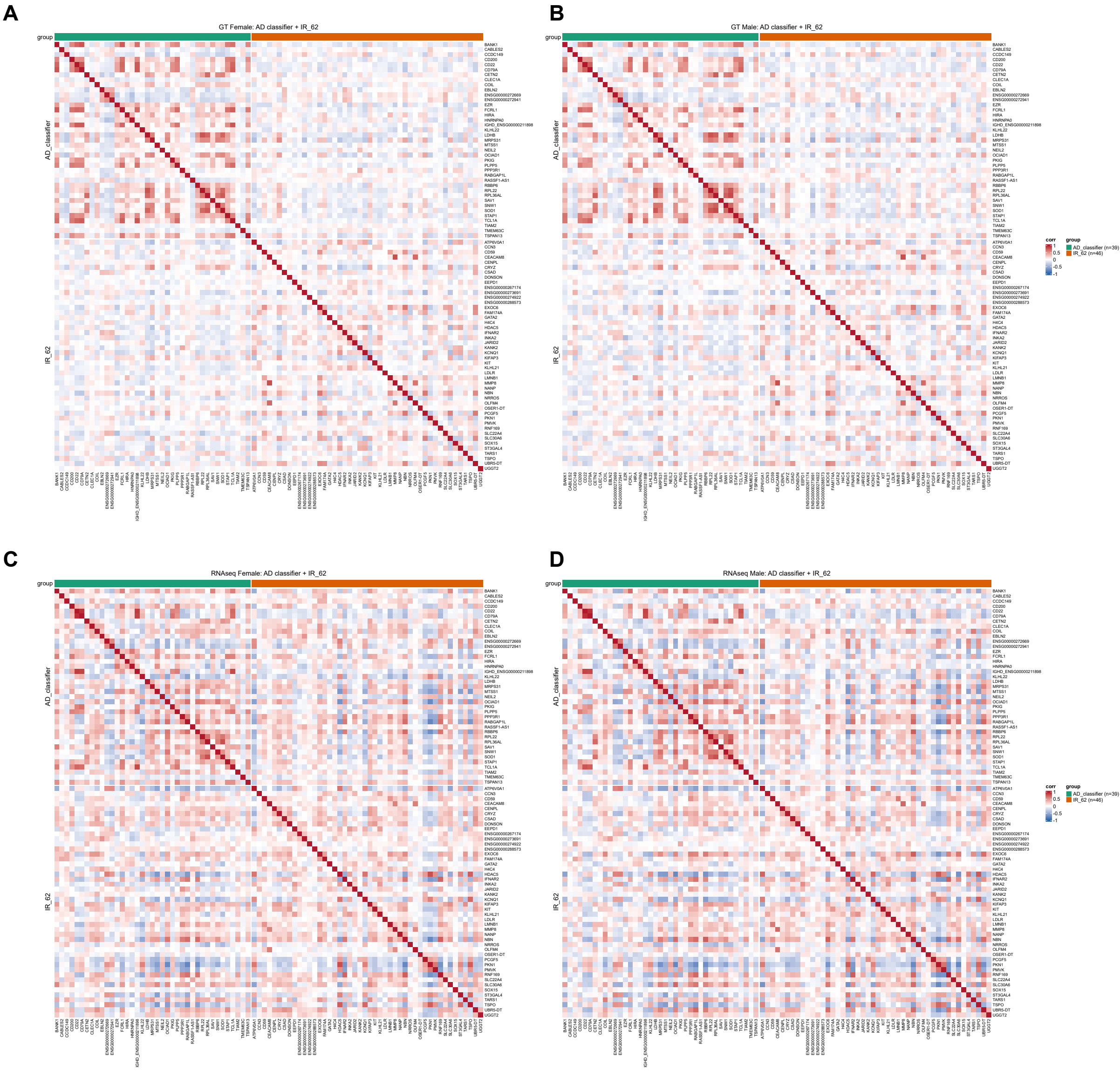


**Figure S15. Correlation heatmap of IR signature and AD classification signature genes in neutrophil-adjusted data.** Pearson correlation coefficients between genes in both the neutrophil-adjusted AD classification array signature and cross-platform IR signature were computed in women and men in the GeneTitan and RNA-seq datasets. Only genes detected in both datasets were included (AD classification signature, n=39; IR signature, n=46). **A:** GeneTitan Female. **B:** GeneTitan Male. **C:** RNAseq Female. **D:** RNAseq Male. Abbreviations: AD, Alzheimer's disease; IR, Insulin resistance; GT, GeneTitan array
