## Supplementary Methods for "Sex and insulin resistance biomarker modelling in a new large-scale Alzheimer’s disease transcriptomic resource"

**Data processing of AddNeuroMed batch 1 Illumina HT12 V3 data (GSE63060)**

48,803 Illumina HT12 V3 probe sequences were converted to FASTA format and aligned to the human reference genome GRCh38 using STAR aligner. A total of 35,456 probes aligned to 97,605 Ensembl transcript IDs (ENSTs). These aligned transcripts were annotated using the BioMart R package to obtain Ensembl Gene IDs (ENSGs), gene symbols, and chromosomal coordinates. Probes mapped to transcripts with chromosomal coordinates within 500 bp of each other were retained. Probes that either mapped to transcripts on different chromosomes or targeted ≥ 2 ENSGs were discarded to reduce cross-hybridisation risks (e.g., pseudogenes, ncRNAs). ENSTs were further filtered to include only those annotated in Affymetrix GeneTitan data (n = 102,673, see methods) to ensure a consistent feature space across platforms for sampling-at-random evaluation (Figure 1E). Following these filtering steps, 23,337 probes remained, representing 66,360 ENSTs. Unnormalised probe intensity values were then quantile normalised using the normalize.quantiles() function from the preprocessCore R package. Normalised intensities were used to calculate the mean and standard deviation (SD) for each probe. Probes were filtered based on linear SD to 16,209 probes (51,267 ENSTs), which were then collapsed to a single row per ENSG, prioritising those with the highest mean intensity, resulting in a dataset of 12,061 probes. This filtering was designed to achieve a similar number of features to our GeneTitan classification dataset (n = 11,607 features) when comparing datasets using sampling-at-random of transcriptomics features (see Figure 1E).
